# Joint associations of device-measured physical activity and sleep duration with incident major adverse cardiovascular events: prospective analysis of the UK Biobank

**DOI:** 10.1101/2025.07.22.25331979

**Authors:** Jason Yun, Laura Brocklebank, Charlie Harper, Aiden Doherty

**Affiliations:** Big Data Institute, Nuffield Department of Population Health, University of Oxford, Old Road Campus, OX3 7LF, UK

## Abstract

**Background:** The interaction between physical activity and sleep with cardiovascular disease remains poorly understood, despite both being key risk factors. This study investigated the independent and joint associations of device-measured physical activity and sleep duration with incident major adverse cardiovascular events (MACE).

**Methods:** Prospective analysis of UK Biobank participants who wore a wrist-based accelerometer for seven days. Open-source machine learning algorithms derived daily step count and overnight sleep duration. The outcome was incident MACE (cardiovascular death, non-fatal myocardial infarction or stroke, or revascularisation procedure), identified through electronic health record linkage. Cox proportional hazards models were used to examine independent and joint associations of median daily step count (low [<7,500], intermediate [7,500-11,000], high [>11,000]) and median overnight sleep duration (short [<6.5 hours], intermediate [6.5-7.5 hours], long [>7.5 hours]) with incident MACE.

**Findings:** Among 88,012 participants (mean age 62·2 years [SD 7·8]), 3,817 were diagnosed with MACE during follow-up (median 7·9 years [IQR 7·3-8·4]). Fewer daily steps and short sleep duration were independently associated with a higher risk of MACE, but there was no evidence of an interaction between step count and sleep duration (*P_interaction_* = 0·42). Compared with the reference group – participants with high step count and intermediate sleep duration – the highest risk of MACE was observed in participants with both low step count and short sleep duration (HR: 1·84, 95% CI: 1·62-2·10).

**Interpretation:** The results of this study show that higher levels of physical activity do not fully attenuate the higher risk of cardiovascular disease associated with short sleep duration, reinforcing the importance of sufficient levels of both physical activity and sleep for the prevention of cardiovascular disease.

**Funding:** Wellcome Trust (223100/Z/21/Z).

**Copyright:** For the purpose of open access, the authors have applied a Creative Commons Attribution (CC BY) licence to any Author Accepted Manuscript version arising from this submission.

**Research in context:** *Evidence before this study:* We searched the Medline and Embase databases in June 2025 for articles published in English from database inception to June 18^th^, 2025. Key search terms included ‘physical activity’, ‘sleep’, and ‘cardiovascular disease’, along with their synonyms, using both MeSH and free-text terms (Table S1). Previous studies have reported mixed evidence for an interaction between physical activity and sleep duration with incident cardiovascular mortality (Table S2 and S3). Some found that higher levels of physical activity attenuated the higher risk associated with abnormal sleep duration, whereas others did not replicate these findings. Many previous studies relied on self-reported measures of physical activity and sleep duration, and reported physical activity metrics that are not easily interpretable. Moreover, by focusing exclusively on cardiovascular mortality, they included relatively few events.

*Added value of this study:* To our knowledge, this prospective cohort study is the first to investigate the joint associations of step count and sleep duration with incident major adverse cardiovascular events. This study used accelerometer data from over 88,000 UK Biobank participants, with the outcome identified through electronic health record linkage. We adjusted for a broad range of key covariates and addressed potential reverse causation by excluding participants with latent or prevalent cardiovascular disease. Although both fewer daily steps and short sleep duration were independently associated with a higher risk of major adverse cardiovascular events, we found no evidence of an interaction between step count and sleep duration.

*Implications of all the available evidence:* Physical activity and sleep are essential components of daily life. They are closely linked within the 24-hour cycle and are both associated with cardiovascular disease. The results of this study show that higher levels of physical activity do not fully attenuate the higher risk of cardiovascular disease associated with short sleep duration, reinforcing the importance of sufficient levels of both physical activity and sleep for the prevention of cardiovascular disease.

## Introduction

Physical activity (PA) and sleep are essential components of daily life, both of which are associated with cardiovascular disease (CVD).^1,2^ Large prospective cohort studies have shown that meeting the World Health Organisation (WHO) recommendation of ≥150 minutes of moderate-intensity aerobic PA per week is associated with a 23% lower risk of CVD mortality.^3^ Sleep duration has been shown to have a J-shaped association with CVD, with both short and long durations linked to higher risk.^4^

PA and sleep are interrelated, with regular PA associated with improved sleep duration and quality.^5^ Furthermore, emerging evidence suggests that higher levels of PA may fully attenuate the higher risk of CVD associated with suboptimal sleep duration.^6,7^ However, this finding has not been replicated in other large prospective cohort studies, resulting in limited understanding of the potential joint associations of PA and sleep with CVD.^8,9^

A major limitation of the current literature is the reliance on self-report questionnaires, which are subject to recall and social desirability biases,^10^ and have been shown to correlate poorly with device-based measures such as polysomnography for sleep and accelerometry for PA.^11,12^ Several studies were statistically underpowered to detect a potential interaction, having ≤1,000 events (Table S2 and S3). Most studies focused exclusively on CVD mortality, limiting understanding of the potential joint associations of PA and sleep with non-fatal cardiovascular events – an important consideration when developing primary prevention strategies. Finally, the use of complex PA metrics, such as metabolic equivalent of task (MET), may hinder the interpretation of findings in a public health context.

Therefore, this study aimed to investigate the independent and joint associations of accelerometer-measured step count and sleep duration with incident major adverse cardiovascular events (MACE) in over 88,000 participants enrolled in the UK Biobank.

## Methods

### Study design, participants, and procedures

This study used data from the UK Biobank, a large prospective cohort study of over 500,000 participants recruited between 2006 and 2010 from across the United Kingdom (excluding Northern Ireland).^13^ Between 2013 and 2015, a subset of 236,519 participants were invited to wear an Axivity AX3 triaxial accelerometer on their dominant wrist continuously for seven days to measure 24-hour movement behaviours, including PA, sedentary behaviour, and sleep.^14^ Of those invited, 106,053 participants agreed to wear the device, and 103,712 provided usable raw acceleration data.

The UK Biobank received ethical approval by the NHS North West Multi-centre Research Ethics Committee (11/NW/0382). Participants gave informed consent to participate in the study before taking part.

### Step count and sleep duration measurements

Step count and sleep duration were derived by applying two open-source machine learning algorithms to the raw acceleration data. These models were developed and evaluated by the Oxford Wearables Group.^15,16^

To derive step count, a hybrid self-supervised machine learning and peak detection algorithm (github.com/OxWearables/stepcount, version 3·7·0) was used, as previously described by Small et al.^15^ Participants were excluded if they did not have sufficient wear time, defined as ≥3 days of data with coverage in every one-hour period of the 24-hour cycle. Non-wear time was defined as unbroken periods of ≥90 minutes during which the SD of acceleration on each axis was <13 m*g*. To address potential diurnal bias in wear time, recording interruptions and non-wear periods were imputed using the average value for the corresponding minute of the day across the remaining valid days.^17^ The primary exposure was median daily step count. Peak 30-minute cadence (a measure of stepping intensity, defined as steps per minute) was calculated by averaging the 30 highest one-minute cadence values per day, then taking the mean across all days in the measurement period.^18^

To derive sleep duration, a self-supervised deep recurrent neural network algorithm (github.com/OxWearables/asleep, version 0·4·12) was used, as previously described by Yuan et al.^16^ Participants were excluded if they did not have sufficient wear time, defined as ≥22 hours per day for ≥3 days (including ≥1 weekend day). Non-wear time was defined identically to the step count algorithm. The primary exposure was median overnight sleep duration, calculated across all noon-to-noon intervals during the measurement period. Sleep efficiency was calculated by dividing overnight sleep duration by the total time spent in bed each day, then averaging this ratio across all valid days.

After excluding participants with insufficient wear time, further exclusions were made if the device could not be calibrated, more than 1% of readings were ‘clipped’ (fell outside ±2 *g*) before or after calibration, average acceleration was implausibly high (>100 m*g*), or step count or sleep duration could not be estimated.

### Outcome ascertainment

The primary outcome was the first occurrence of a MACE event, defined as death from any cardiovascular cause, non-fatal myocardial infarction or stroke, or revascularisation procedure. MACE events were identified through linkage to National Health Service (NHS) Digital for England, Secure Anonymised Information Linkage (SAIL) Databank for Wales, and the NHS Central Register for Scotland (Table S4). To identify incident cases of MACE, participants with a prior self-reported or hospital-recorded diagnosis were excluded from the analysis.

### Covariates

Potential confounders and mediators were selected a priori using a causal diagram based on the current literature (Figure S1). These variables were derived from self-report (demographic, socioeconomic, and lifestyle factors), physical measurements (body mass index [BMI] and blood pressure), or blood samples (glycated haemoglobin [HbA1c] and total cholesterol) (Table S5).

### Statistical analysis

To aid interpretability, median daily step count was initially divided into tertiles and then reclassified by rounding the boundaries to the nearest 500 steps (low [<7,500], intermediate [7,500–11,000], high [>11,000]). Median overnight sleep duration was categorised similarly, with tertile boundaries rounded to the nearest 0·5 hours (short [<6·5 hours], intermediate [6·5-7·5 hours], long [>7·5 hours]). This approach was adopted due to the lack of consensus in the current literature on accelerometer-based cut-points for step count and sleep duration, with existing guidelines largely derived from self-reported data.^19,20^

Baseline participant characteristics were reported as mean and standard deviation (SD) for normally distributed continuous variables, median and interquartile range (IQR) for non-normally distributed continuous variables, and number of participants and percentage (%) for categorical variables. Univariable associations between each covariate and the exposures (step count and sleep duration) were assessed using the Pearson χ² test for categorical covariates, the one-way Analysis of Variance (ANOVA) test for normally distributed continuous covariates, and the Kruskal-Wallis test for non-normally distributed continuous covariates. Associations between each covariate and the outcome (incident MACE) were assessed using univariable Cox proportional hazards models.

Multivariable Cox proportional hazards models were employed to examine the independent associations of step count and sleep duration with incident MACE,^21^ using age as the timescale consistent with previous studies.^15,16^ Sequential adjustments for potential confounders were performed in the following order: ethnicity, education, Townsend Deprivation Index (TDI), smoking status, alcohol intake, processed meat intake, and finally step count or sleep duration (depending on the exposure). Both minimally adjusted and maximally adjusted hazard ratios (HR) with their respective 95% confidence intervals (CIs) were reported in the main text. Floating absolute risks were also calculated to provide risk estimates and 95% CIs for each group independently, allowing direct comparisons between groups. These were used where appropriate to display 95% CIs in the figures.^22^ In addition to the maximally adjusted model, we further adjusted for each of the following potential mediators individually: BMI, HbA1c, blood pressure, and total cholesterol.

The potential interaction between step count and sleep duration with incident MACE was explored by stratifying participants into nine mutually exclusive groups, with group-specific HRs reported (Table S6). Participants with high step count and intermediate sleep duration served as the reference group. To assess evidence of an interaction, a likelihood ratio (LR) test was performed comparing the maximally adjusted model with and without an interaction term between step count and sleep duration.

### Sensitivity analysis

To examine potential reverse causation, participants diagnosed with MACE within the first two, four, and six years of follow-up were sequentially excluded. Additional analyses excluded participants with a prior self-reported or hospital-recorded diagnosis of any CVD or cancer, identified using the relevant medical classification codes (Table S4). To assess the impact of sleep duration misclassification, participants with factors that could disrupt sleep were excluded, including self-reported shift workers, those with a prior diagnosis of sleep apnoea or restless leg syndrome, and those whose accelerometer wear period overlapped with daylight saving time changes. Finally, adjustments for sleep efficiency and step cadence were performed to assess the potential confounding effects of sleep quality and stepping intensity on the observed associations.

All analyses were conducted using STATA (version 18·0), and evidence of suggestive statistical significance was defined as *p* <0·05. Results have been reported according to the STROBE guidelines (Table S7).^23^

## Results

### Baseline characteristics

Of the 103,712 participants with usable raw acceleration data, 52 were excluded due to withdrawal or loss of linked electronic health record follow-up before the accelerometry sub-study. Among the remaining 103,660, exclusions included 9,072 for invalid accelerometer data, 4,463 for prior MACE, and 2,113 for missing covariates, resulting in a final analytic sample of 88,012 participants (Figure 1).

**Figure 1.**
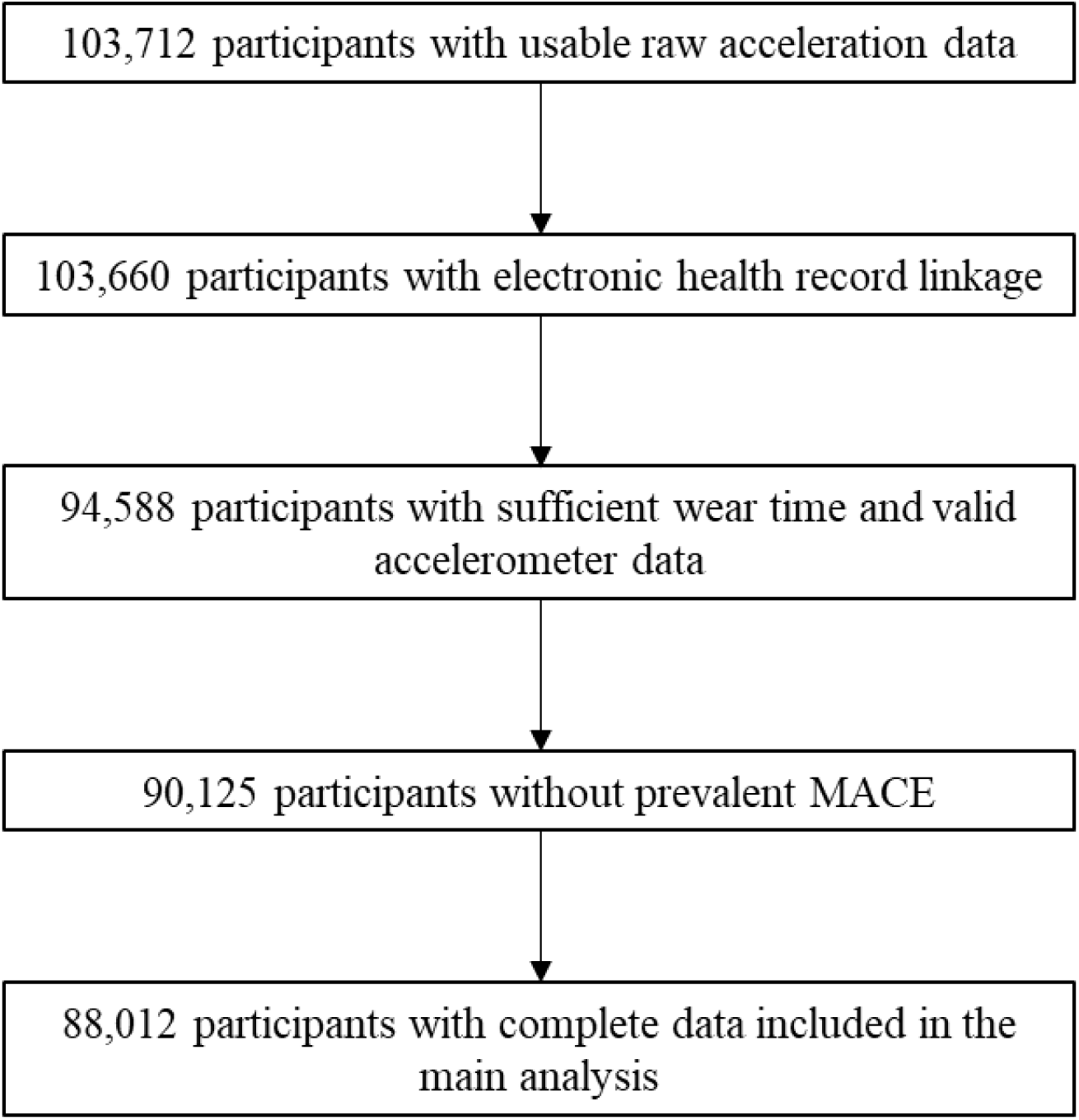
Flow diagram for participants included in the analysis of the independent and joint associations of accelerometer-measured step count and sleep duration with incident major adverse cardiovascular events (MACE).

Table 1 presents the baseline characteristics of the final analytic sample, grouped by median daily step count. Among the participants, 58·0% (N: 51,045) were women, with a mean age of 62·2 years (SD: 7·8) at the end of accelerometer wear. Most were white (96·9%), from the least deprived TDI quintile (50·7%), and had a high level of education (43·9%). Compared with the high step count group, the low step count group had fewer participants with a high level of education, more current smokers, more obese participants, more long sleepers, and fewer short sleepers.

**Table 1.**
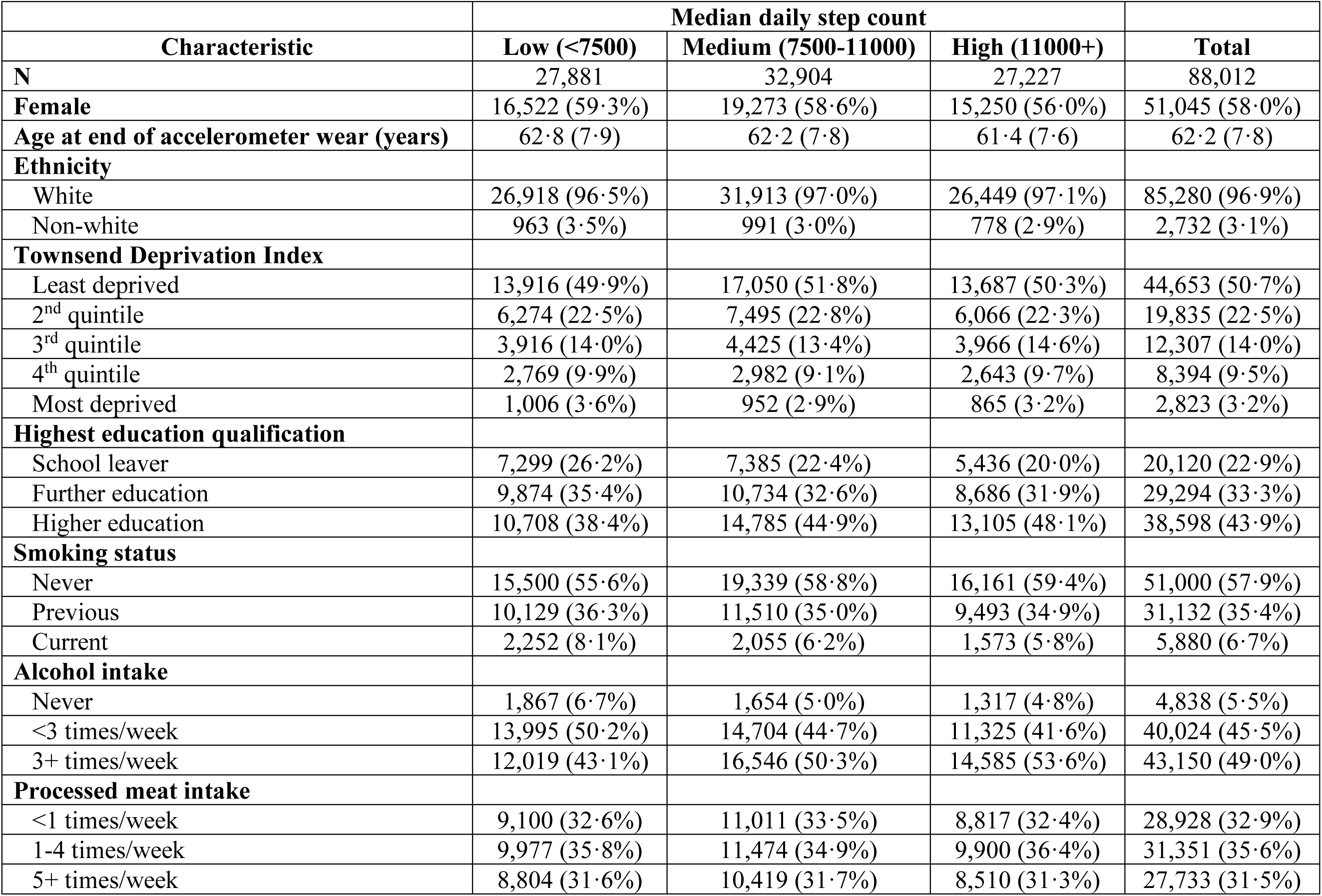
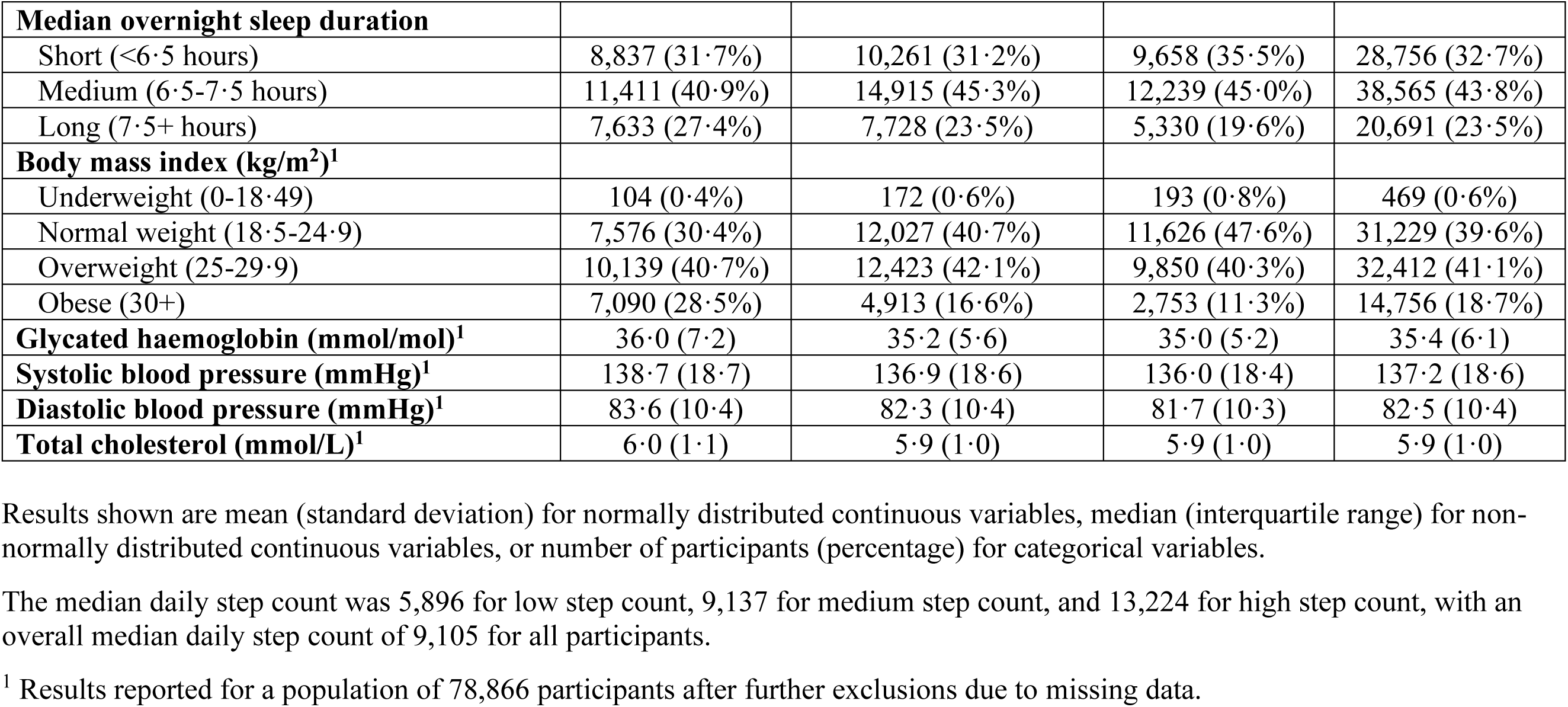
Baseline characteristics of study participants by step count.

| Characteristic | Median daily step count |  |  | Total |
| --- | --- | --- | --- | --- |
|  | Low (<7500) | Medium (7500-11000) | High (11000+) |  |
| **N** | 27,881 | 32,904 | 27,227 | 88,012 |
| **Female** | 16,522 (59.3%) | 19,273 (58.6%) | 15,250 (56.0%) | 51,045 (58.0%) |
| **Age at end of accelerometer wear (years)** | 62.8 (7.9) | 62.2 (7.8) | 61.4 (7.6) | 62.2 (7.8) |
| **Ethnicity** |  |  |  |  |
| White | 26,918 (96.5%) | 31,913 (97.0%) | 26,449 (97.1%) | 85,280 (96.9%) |
| Non-white | 963 (3.5%) | 991 (3.0%) | 778 (2.9%) | 2,732 (3.1%) |
| **Townsend Deprivation Index** |  |  |  |  |
| Least deprived | 13,916 (49.9%) | 17,050 (51.8%) | 13,687 (50.3%) | 44,653 (50.7%) |
| 2<sup>nd</sup> quintile | 6,274 (22.5%) | 7,495 (22.8%) | 6,066 (22.3%) | 19,835 (22.5%) |
| 3<sup>rd</sup> quintile | 3,916 (14.0%) | 4,425 (13.4%) | 3,966 (14.6%) | 12,307 (14.0%) |
| 4<sup>th</sup> quintile | 2,769 (9.9%) | 2,982 (9.1%) | 2,643 (9.7%) | 8,394 (9.5%) |
| Most deprived | 1,006 (3.6%) | 952 (2.9%) | 865 (3.2%) | 2,823 (3.2%) |
| **Highest education qualification** |  |  |  |  |
| School leaver | 7,299 (26.2%) | 7,385 (22.4%) | 5,436 (20.0%) | 20,120 (22.9%) |
| Further education | 9,874 (35.4%) | 10,734 (32.6%) | 8,686 (31.9%) | 29,294 (33.3%) |
| Higher education | 10,708 (38.4%) | 14,785 (44.9%) | 13,105 (48.1%) | 38,598 (43.9%) |
| **Smoking status** |  |  |  |  |
| Never | 15,500 (55.6%) | 19,339 (58.8%) | 16,161 (59.4%) | 51,000 (57.9%) |
| Previous | 10,129 (36.3%) | 11,510 (35.0%) | 9,493 (34.9%) | 31,132 (35.4%) |
| Current | 2,252 (8.1%) | 2,055 (6.2%) | 1,573 (5.8%) | 5,880 (6.7%) |
| **Alcohol intake** |  |  |  |  |
| Never | 1,867 (6.7%) | 1,654 (5.0%) | 1,317 (4.8%) | 4,838 (5.5%) |
| <3 times/week | 13,995 (50.2%) | 14,704 (44.7%) | 11,325 (41.6%) | 40,024 (45.5%) |
| 3+ times/week | 12,019 (43.1%) | 16,546 (50.3%) | 14,585 (53.6%) | 43,150 (49.0%) |
| **Processed meat intake** |  |  |  |  |
| <1 times/week | 9,100 (32.6%) | 11,011 (33.5%) | 8,817 (32.4%) | 28,928 (32.9%) |
| 1-4 times/week | 9,977 (35.8%) | 11,474 (34.9%) | 9,900 (36.4%) | 31,351 (35.6%) |
| 5+ times/week | 8,804 (31.6%) | 10,419 (31.7%) | 8,510 (31.3%) | 27,733 (31.5%) |
| **Median overnight sleep duration** |  |  |  |  |
| Short (<6.5 hours) | 8,837 (31.7%) | 10,261 (31.2%) | 9,658 (35.5%) | 28,756 (32.7%) |
| Medium (6.5-7.5 hours) | 11,411 (40.9%) | 14,915 (45.3%) | 12,239 (45.0%) | 38,565 (43.8%) |
| Long (7.5+ hours) | 7,633 (27.4%) | 7,728 (23.5%) | 5,330 (19.6%) | 20,691 (23.5%) |
| **Body mass index (kg/m<sup>2</sup>)<sup>1</sup>** |  |  |  |  |
| Underweight (0-18.49) | 104 (0.4%) | 172 (0.6%) | 193 (0.8%) | 469 (0.6%) |
| Normal weight (18.5-24.9) | 7,576 (30.4%) | 12,027 (40.7%) | 11,626 (47.6%) | 31,229 (39.6%) |
| Overweight (25-29.9) | 10,139 (40.7%) | 12,423 (42.1%) | 9,850 (40.3%) | 32,412 (41.1%) |
| Obese (30+) | 7,090 (28.5%) | 4,913 (16.6%) | 2,753 (11.3%) | 14,756 (18.7%) |
| **Glycated haemoglobin (mmol/mol)<sup>1</sup>** | 36.0 (7.2) | 35.2 (5.6) | 35.0 (5.2) | 35.4 (6.1) |
| **Systolic blood pressure (mmHg)<sup>1</sup>** | 138.7 (18.7) | 136.9 (18.6) | 136.0 (18.4) | 137.2 (18.6) |
| **Diastolic blood pressure (mmHg)<sup>1</sup>** | 83.6 (10.4) | 82.3 (10.4) | 81.7 (10.3) | 82.5 (10.4) |
| **Total cholesterol (mmol/L)<sup>1</sup>** | 6.0 (1.1) | 5.9 (1.0) | 5.9 (1.0) | 5.9 (1.0) |

Results shown are mean (standard deviation) for normally distributed continuous variables, median (interquartile range) for non-normally distributed continuous variables, or number of participants (percentage) for categorical variables.
The median daily step count was 5,896 for low step count, 9,137 for medium step count, and 13,224 for high step count, with an overall median daily step count of 9,105 for all participants.
<sup>1</sup> Results reported for a population of 78,866 participants after further exclusions due to missing data.

### Independent association of step count and incident major adverse cardiovascular events

Among the 88,012 participants, 3,817 cases of MACE were recorded over 696,294 person-years of follow-up (median follow-up: 7·9 years, IQR: 7·3-8·4). Fewer daily steps were associated with a higher risk of MACE (Figure 2A). In the maximally adjusted model, participants in the low step count group had a 52% higher risk (HR: 1·52, 95% CI: 1·40-1·65; Figure S2A), and those in the intermediate group had a 24% higher risk (HR: 1·24, 95% CI: 1·14-1·35; Figure S2B), compared with the high step count reference group. BMI modestly attenuated the associations between step count and incident MACE (low: HR 1·42, 95% CI 1·30-1·55; intermediate: HR 1·21, 95% CI 1·11-1·32), whereas HbA1c, blood pressure, and total cholesterol had minimal impact (Figures S3A and S3B).

**Figure 2.**
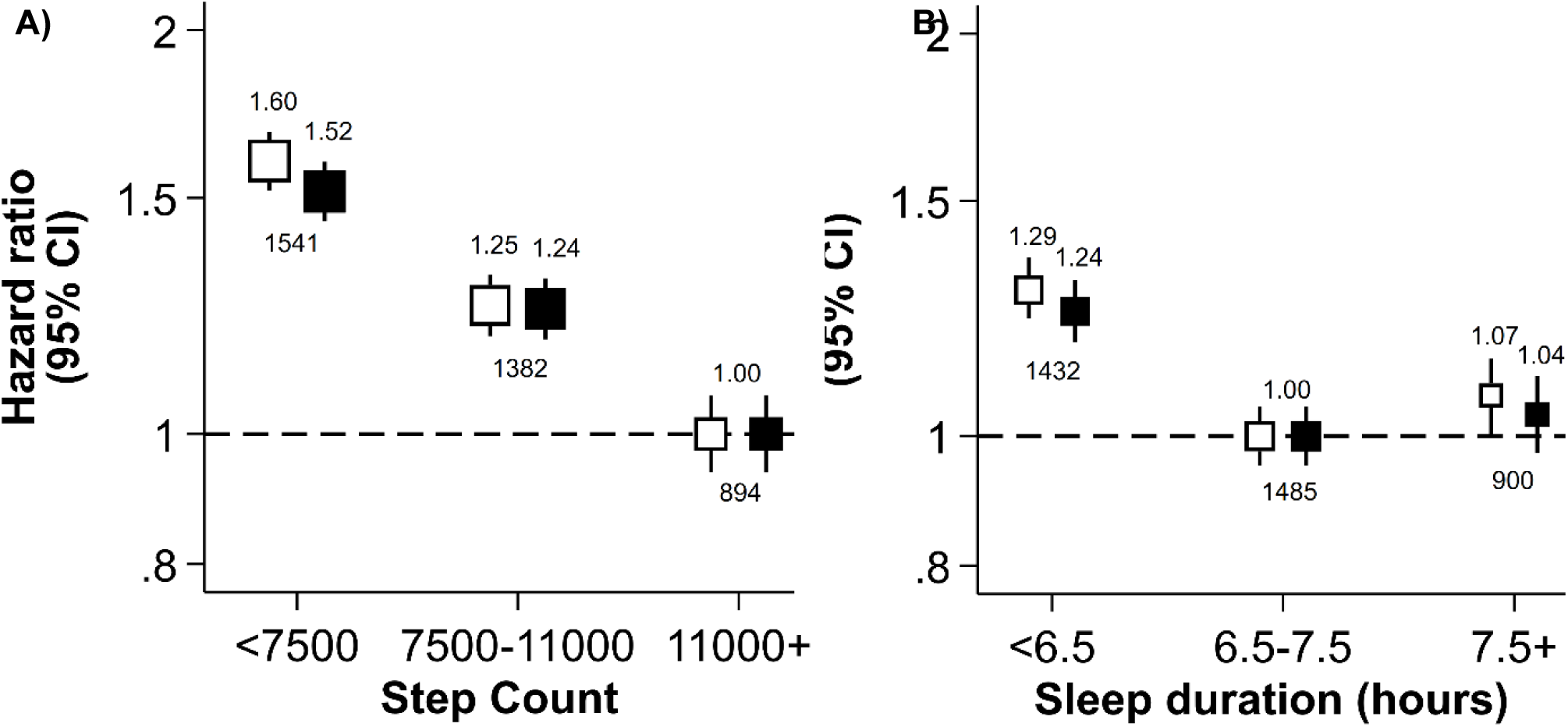
Independent associations of step count (A) and sleep duration (B) with incident major adverse cardiovascular events (MACE). HRs from both the minimally adjusted model (white boxes; adjusted for age and sex) and the maximally adjusted model (black boxes; additionally adjusted for ethnicity, education, TDI, smoking status, alcohol intake, processed meat intake, and sleep duration or step count) have been depicted for each participant group according to the following classifications for median daily step count (low [<7500], intermediate [7500-11000], high [11000+; reference) and median overnight sleep duration (short [<6.5 hours], intermediate [6.5-7.5 hours; reference], long [7.5+ hours]). The vertical lines represent 95% CIs based on floating absolute risks. The size of each box is relative to the amount of statistical information available. Numbers above the boxes indicate the HRs, while numbers below indicate the total number of events in each group. The HRs for the y-axis have been plotted on a log scale. HR, hazard ratio; TDI, Townsend Deprivation Index; 95% CI, 95% confidence interval.

Excluding MACE cases within the first two, four, and six years progressively attenuated the associations (Figures S4A and S4B). After excluding the first six years, the low step count group had a 22% higher risk than the reference group (HR: 1·22, 95% CI: 1·05-1·42), while the association for the intermediate group was no longer suggestive of statistical significance (HR: 1·14, 95% CI: 0·98-1·32). Excluding those with prior CVD or cancer attenuated the association for low step count (HR: 1·35, 95% CI: 1·18-1·54; Figure S5A), but not for intermediate step count (Figure S5B). Excluding participants with factors that could disrupt sleep did not substantially alter the associations (Figures S6A and S6B). Finally, adjustment for step cadence, but not sleep efficiency, attenuated the associations (Figures S7A and S7B).

### Independent association of sleep duration and incident major adverse cardiovascular events

Short sleep duration was associated with a higher risk of MACE (Figure 2B). In the maximally adjusted model, participants with short sleep duration had a 24% higher risk compared with the intermediate sleep duration reference group (HR: 1·24, 95% CI: 1·15-1·33; Figure S2C). In contrast, the risk of MACE among participants with long sleep duration was not significantly different from the reference group (HR: 1·04, 95% CI: 0·96-1·13; Figure S2D). BMI modestly attenuated the association for short sleep duration (HR: 1·21, 95% CI: 1·12-1·30), whereas HbA1c, blood pressure, and total cholesterol had minimal impact (Figure S3C). The non-significant association for long sleep duration persisted across all mediator adjustments (Figure S3D).

Excluding participants with prior CVD or cancer fully attenuated the association between short sleep duration and incident MACE (HR: 1·10, 95% CI: 0·97-1·23; Figure S5C). The other sensitivity analyses did not substantially alter the risk estimate (Figures S4C, S6C, and S7C). The non-significant association for long sleep duration persisted across all sensitivity analyses (Figures S4D-S7D).

### Joint associations of step count and sleep duration with incident major adverse cardiovascular events

Overall, there was no evidence of an interaction between step count and sleep duration with incident MACE (*P_interaction_* = 0.42). Compared with the reference group – high step count and intermediate sleep duration – all other groups had a higher risk of MACE (Figure 3 and Table S6), except participants with high step count and long sleep duration (HR: 0·97, 95% CI: 0·83-1·13). The highest risk was among those with both low step count and short sleep duration (HR: 1·84, 95% CI: 1·70-2·00). Including all potential mediators (BMI, HbA1c, blood pressure, and total cholesterol) did not substantially alter the interaction or joint risk estimates (*P_interaction_* = 0.54; Table S6 and Figure S8).

**Figure 3.**
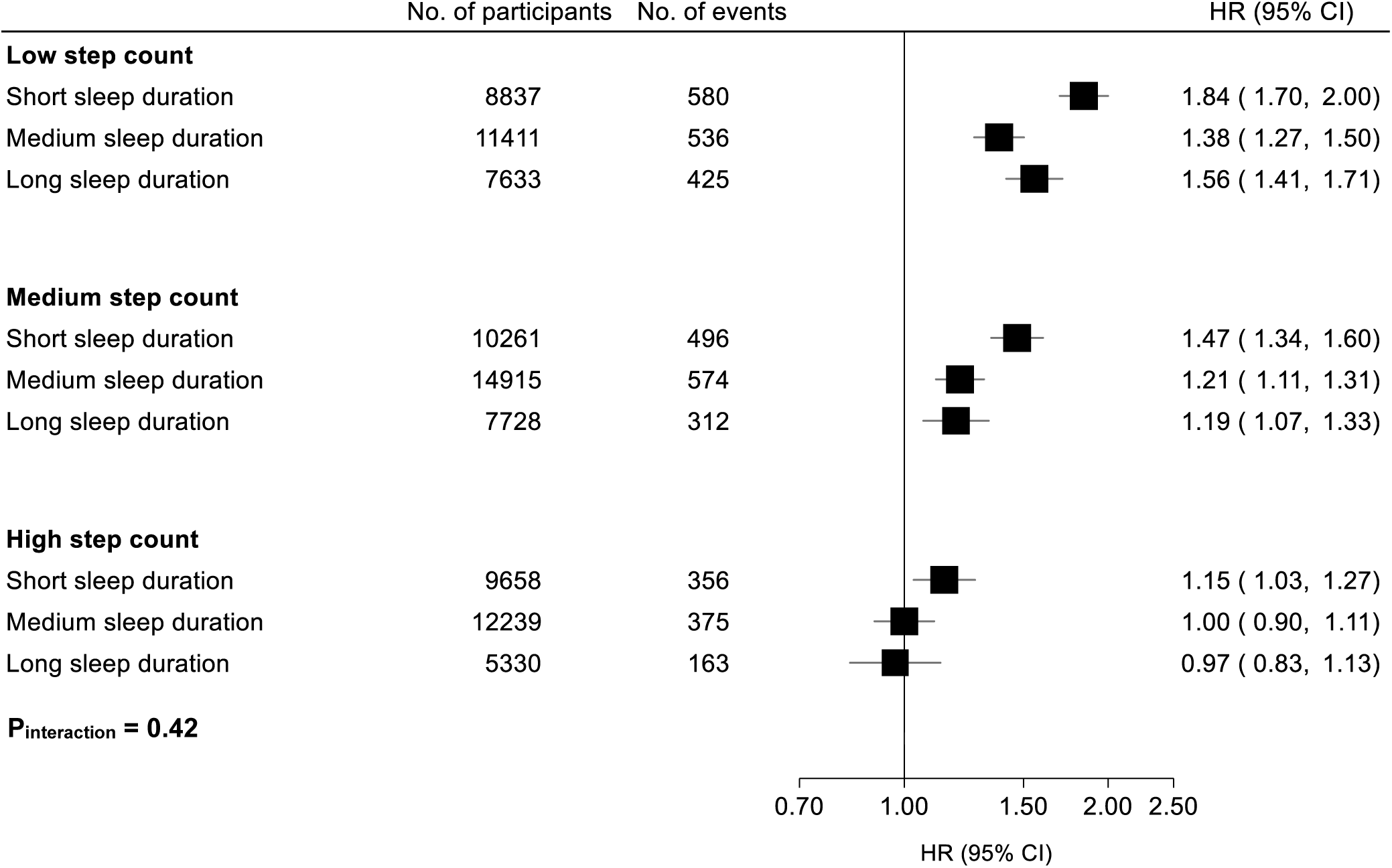
Joint associations of step count and sleep duration with incident major adverse cardiovascular events (MACE). Participants were stratified into nine mutually exclusive groups using the following classifications for median daily step count (low [<7500], intermediate [7500-11000], high [11000+; reference) and median overnight sleep duration (short [<6.5 hours], intermediate [6.5-7.5 hours; reference], long [7.5+ hours]). HRs from the maximally adjusted model (adjusted for age, sex, ethnicity, education, TDI, smoking status, alcohol intake, and processed meat intake) have been depicted for each participant group. The *p*-value for interaction was derived from a likelihood ratio test comparing the model with and without an interaction term between step count and sleep duration. The horizontal lines represent 95% CIs based on floating absolute risks. The size of each box is relative to the amount of statistical information available. The HRs for the x-axis have been plotted on a log scale. HR, hazard ratio; TDI, Townsend Deprivation Index; 95% CI, 95% confidence interval.

Excluding participants with prior CVD or cancer substantially attenuated the joint risk estimates, with only a few combinations remaining associated with a higher risk of MACE – specifically, intermediate step count with short sleep duration, and low step count with either short or long sleep duration (Table S6 and Figure S9). Excluding MACE cases within the first four years (Table S6 and Figure S10) and adjusting for step cadence and sleep efficiency (Table S6 and Figure S11) modestly attenuated the risk estimates, while excluding those with sleep-disrupting factors did not (Table S6 and Figure S12). Across all sensitivity analyses, there was no evidence of an interaction between step count and sleep duration with incident MACE.

## Discussion

In this study of over 88,000 UK Biobank participants with accelerometer data, we found no evidence of an interaction between step count and sleep duration with incident MACE. However, there was a strong independent inverse association between step count and incident MACE, with participants in the low step count group having a ∼40% higher risk compared with those in the high step count group. We also observed an independent L-shaped association between sleep duration and incident MACE, with participants in the short sleep duration group having a ∼20% higher risk compared with those with intermediate sleep duration. These findings highlight the importance of maintaining sufficient levels of both PA and sleep for the prevention of CVD, which could help inform future public health guidelines.

The lack of interaction in this study contrasts with findings from some previous studies using both self-reported and device-based data. For example, using self-reported data from over 340,000 participants in Taiwan, Chen et al. reported that those with low PA and long sleep duration had a 66% higher risk of CVD mortality compared with the reference group – participants with high PA and intermediate sleep duration (HR: 1·66, 95% CI: 1·43-1·92). They also found that high PA fully attenuated the elevated risk associated with long sleep duration (HR: 1·02, 95% CI: 0·81-1·29).^6^ In contrast, we found no association between long sleep duration and incident MACE. As Chen et al.’s study relied solely on self-reported measures, misclassification bias may have contributed to this discrepancy. For example, participants with poorer cardiovascular health may have overestimated their sleep duration.^24^

Liang et al. also reported an interaction between device-based PA and sleep duration with the risk of CVD mortality. Participants with both low PA and short sleep duration had nearly a four-fold higher risk than those with high PA and intermediate sleep duration (HR: 3·93, 95% CI: 2·90-5·32), and higher PA fully attenuated the elevated risk associated with short sleep duration (HR:1·40, 95% CI: 0·88-2·25).^7^ We found no evidence of an interaction between PA and sleep duration with incident MACE. However, similar to Liang et al., we observed that participants with both low PA and short sleep duration had the highest risk of MACE, although the association was weaker in our study (HR: 1·84, 95% CI: 1·62-2·10). This difference likely reflects our inclusion of non-fatal CVD events and revascularisation procedures, while more severe outcomes such as CVD mortality tend to show stronger associations with PA and sleep. However, our broader MACE definition resulted in more cases (N: 3,817) than Liang et al. (N: 1,074), reducing random error – an important consideration when stratifying by multiple groups to examine interaction effects.

The independent association between step count and incident MACE in our study aligns with the existing literature. For example, a meta-analysis by Stens et al., using device-based step count data from over 85,000 participants across four prospective cohort studies, found that those in the low step count group had more than twice the risk of CVD compared with the high step count group (HR: 2·38, 95% CI: 1·88-3·03).^20^ Our more conservative estimate may reflect the higher median daily step count in our low step count group (5,896 steps) compared with theirs (2,022 steps), as lower step counts tend to show stronger associations with incident CVD.

Our findings on the independent association between sleep duration and incident MACE differ from those of previous studies. For example, a large meta-analysis of over three million participants across 74 observational studies reported a J-shaped association, with both short (<6 hours) and long (>9 hours) sleep durations associated with a higher risk of CVD mortality compared with intermediate sleep duration (7 hours).^19^ In contrast, we found that only short sleep duration was associated with a higher risk of MACE. Besides misclassification bias from self-reported sleep duration, this discrepancy may stem from some of the meta-analysis studies not excluding participants with latent or prevalent CVD, potentially introducing reverse causation bias where long sleep duration reflects underlying disease.^25^

Our findings suggest reverse causality may influence both the independent and joint associations of step count and sleep duration with incident MACE. Excluding the first six years of follow-up notably attenuated the risks associated with fewer daily steps, but not short sleep duration, possibly because latent CVD more strongly affects PA than sleep duration.^24,26^ However, excluding participants with prior CVD or cancer attenuated the associations for both exposures, likely due to the removal of participants with more chronic conditions, such as heart failure, which impact both PA and sleep duration.^25^

Strengths of our study include the large number of MACE events, which provided sufficient statistical power even after categorising participants into nine groups, as well as the small proportion of missing covariate data (∼2·0%) and loss to follow-up (<0·01%). We used open-source machine learning algorithms to derive step count and sleep duration from raw acceleration data, which have shown greater reliability and validity than self-reported measures.^15,16^ PA was assessed using step count – a clinically interpretable and widely adopted metric^20^ – enhancing the applicability of our findings to health promotion strategies. We also adjusted for a broad range of key covariates, allowing us to assess their potential confounding and mediating effects on the observed associations.

However, several limitations of our study should be acknowledged. The measurement period was limited to a maximum of seven days per participant, which may not fully capture habitual PA and sleep patterns. This could lead to regression dilution bias and an underestimation of the true associations.^27^ Most covariates were measured at study entry, several years before accelerometer wear, potentially introducing misclassification bias and residual confounding. The external validity of our findings may be limited by the low representativeness of UK Biobank participants, who have been shown to be more physically active, wealthier, and healthier than the general population.^28^ Additionally, very low step counts and very long sleep durations were rare in our sample, limiting statistical power to examine their associations with incident MACE. Finally, as with all observational studies, causal inferences cannot be made.

In summary, fewer daily steps and short sleep duration were independently associated with a higher risk of MACE, with no evidence of an interaction between the two. Further research using repeated, longer-term measurements, extended follow-up to mitigate reverse causation, and replication in other cohorts is warranted.

## Data Availability

Data may be obtained from a third party and are not publicly available. This research was conducted using the UK Biobank Resource under Application Number 59070. Data are accessible through the UK Biobank following an application process and approval from the UK Biobank Research Ethics Committee.

## Contributors

Concept and design: JY, CH, and AD. Data access and verification: JY and CH. Statistical analysis: JY. Data interpretation: all authors. Drafting of the manuscript: LB and JY. Critical revision of the manuscript: all authors. All authors had full access to all the data in the study and had final responsibility for the decision to submit for publication.

## Acknowledgements

AD’s research team is supported by a range of grants from the Wellcome Trust (223100/Z/21/Z, 227093/Z/23/Z), Novo Nordisk, Swiss Re, Boehringer Ingelheim, National Institutes of Health’s Oxford-Cambridge Scholars Program, EPSRC Centre for Doctoral Training in Health Data Science (EP/S02428X/1), British Heart Foundation Centre of Research Excellence (RE/18/3/34214), and funding administered by the Danish National Research Foundation in support of the Pioneer Centre for SMARTbiomed. LB is supported by the Wellcome Trust, and CH was supported by Swiss Re.

## References

1 Fiuza-Luces C, Santos-Lozano A, Joyner M, et al. Exercise benefits in cardiovascular disease: beyond attenuation of traditional risk factors. Nature Reviews Cardiology 2018; 15(12): 731–43

2 Wang T, Zhao Z, Yu X, et al. Age-specific modifiable risk factor profiles for cardiovascular disease and all-cause mortality: a nationwide, population-based, prospective cohort study. The Lancet Regional Health – Western Pacific 2021; 17

3 Wahid A, Manek N, Nichols M, et al. Quantifying the Association Between Physical Activity and Cardiovascular Disease and Diabetes: A Systematic Review and Meta-Analysis. J Am Heart Assoc 2016; 5(9)

4 Wang S, Li Z, Wang X, et al. Associations between sleep duration and cardiovascular diseases: A meta-review and meta-analysis of observational and Mendelian randomization studies. Front Cardiovasc Med 2022; 9: 930000

5 Kredlow MA, Capozzoli MC, Hearon BA, Calkins AW, Otto MW. The effects of physical activity on sleep: a meta-analytic review. Journal of Behavioral Medicine 2015; 38(3): 427–49

6 Chen LJ, Hamer M, Lai YJ, Huang BH, Ku PW, Stamatakis E. Can physical activity eliminate the mortality risk associated with poor sleep? A 15-year follow-up of 341,248 MJ Cohort participants. J Sport Health Sci 2022; 11(5): 596–604

7 Liang YY, Feng H, Chen Y, et al. Joint association of physical activity and sleep duration with risk of all-cause and cause-specific mortality: a population-based cohort study using accelerometry. Eur J Prev Cardiol 2023; 30(9): 832–43

8 Xiao Q, Keadle SK, Hollenbeck AR, Matthews CE. Sleep duration and total and cause-specific mortality in a large US cohort: interrelationships with physical activity, sedentary behavior, and body mass index. Am J Epidemiol 2014; 180(10): 997–1006

9 Wang W, Yang J, Wang K, et al. Association between self-reported sleep duration, physical activity and the risk of all cause and cardiovascular diseases mortality from the NHANES database. BMC Cardiovasc Disord 2023; 23(1): 467

10 Althubaiti A. Information bias in health research: definition, pitfalls, and adjustment methods. J Multidiscip Healthc 2016; 9: 211–7

11 Lauderdale DS, Knutson KL, Yan LL, Liu K, Rathouz PJ. Self-reported and measured sleep duration: how similar are they? Epidemiology 2008; 19(6): 838–45

12 Skender S, Ose J, Chang-Claude J, et al. Accelerometry and physical activity questionnaires - a systematic review. BMC Public Health 2016; 16(1): 515

13 Sudlow C, Gallacher J, Allen N, et al. UK biobank: an open access resource for identifying the causes of a wide range of complex diseases of middle and old age. PLoS Med 2015; 12(3): e1001779

14 Doherty A, Jackson D, Hammerla N, et al. Large Scale Population Assessment of Physical Activity Using Wrist Worn Accelerometers: The UK Biobank Study. PLOS ONE 2017; 12(2): e0169649

15 Small SR, Chan S, Walmsley R, et al. Self-Supervised Machine Learning to Characterise Step Counts from Wrist-Worn Accelerometers in the UK Biobank. Med Sci Sports Exerc 2024; 56(10): 1945–53

16 Yuan H, Plekhanova T, Walmsley R, et al. Self-supervised learning of accelerometer data provides new insights for sleep and its association with mortality. npj Digital Medicine 2024; 7(1): 86

17 van Hees VT, Fang Z, Langford J, et al. Autocalibration of accelerometer data for free-living physical activity assessment using local gravity and temperature: an evaluation on four continents. Journal of Applied Physiology 2014; 117(7): 738–44

18 Saint-Maurice PF, Troiano RP, Bassett DR, et al. Association of Daily Step Count and Step Intensity With Mortality Among US Adults. JAMA 2020; 323(12): 1151–60

19 Kwok CS, Kontopantelis E, Kuligowski G, et al. Self-Reported Sleep Duration and Quality and Cardiovascular Disease and Mortality: A Dose-Response Meta-Analysis. Journal of the American Heart Association 2018; 7(15): e008552

20 Stens NA, Bakker EA, Mañas A, et al. Relationship of Daily Step Counts to All-Cause Mortality and Cardiovascular Events. J Am Coll Cardiol 2023; 82(15): 1483–94

21 Cox DR. Regression Models and Life-Tables. Journal of the Royal Statistical Society: Series B (Methodological) 1972; 34(2): 187–202

22 Easton DF, Peto J, Babiker AG. Floating absolute risk: an alternative to relative risk in survival and case-control analysis avoiding an arbitrary reference group. Statistics in medicine 1991; 10(7): 1025–35

23 von Elm E, Altman DG, Egger M, Pocock SJ, Gøtzsche PC, Vandenbroucke JP. The Strengthening the Reporting of Observational Studies in Epidemiology (STROBE) statement: guidelines for reporting observational studies. J Clin Epidemiol 2008; 61(4): 344–9

24 Suzuki K, Miyamoto M, Hirata K. Sleep disorders in the elderly: Diagnosis and management. J Gen Fam Med 2017; 18(2): 61–71

25 Ziegler KA, Ahles A, Dueck A, et al. Immune-mediated denervation of the pineal gland underlies sleep disturbance in cardiac disease. Science 2023; 381(6655): 285–90

26 Strain T, Wijndaele K, Sharp SJ, Dempsey PC, Wareham N, Brage S. Impact of follow-up time and analytical approaches to account for reverse causality on the association between physical activity and health outcomes in UK Biobank. International Journal of Epidemiology 2020; 49(1): 162–72

27 Saint-Maurice PF, Sampson JN, Keadle SK, Willis EA, Troiano RP, Matthews CE. Reproducibility of Accelerometer and Posture-derived Measures of Physical Activity. Med Sci Sports Exerc 2020; 52(4): 876–83

28 Fry A, Littlejohns TJ, Sudlow C, et al. Comparison of Sociodemographic and Health-Related Characteristics of UK Biobank Participants With Those of the General Population. Am J Epidemiol 2017; 186(9): 1026–3

