## Supplementary Material for "Joint associations of device-measured physical activity and sleep duration with incident major adverse cardiovascular events: prospective analysis of the UK Biobank"

### Table of contents


| | |
|---|---|
| Page 3 | Table S1. Search terms and results from literature review. |
| Page 4 | Table S2. Summary of study characteristics for articles included in literature review. |
| Pages 5-7 | Table S3. Summary of main results for articles included in literature review. |
| Page 8 | Table S4. Medical classification codes for incident major adverse cardiovascular events (MACE; ICD-10), previous disease (ICD-10), and revascularisation procedures (OPCS-4). |
| Page 9 | Table S5. UK Biobank fields for all variables used. |
| Pages 10-11 | Table S6. Hazard ratios and their respective conventional and floating absolute risk confidence intervals for the joint associations of step count and sleep duration with incident major adverse cardiovascular events (MACE). |
| Pages 12-13 | Table S7. STROBE statement checklist. |
| Page 14 | Figure S1. Causal diagram depicting the mechanistic pathways between physical activity, sleep, key covariates, and cardiovascular disease. |
| Pages 15-16 | Figure S2. Independent associations of step count (A and B) and sleep duration (C and D) with incident major adverse cardiovascular events (MACE), with sequential adjustments for potential confounders. |
| Pages 17-18 | Figure S3. Independent associations of step count (A and B) and sleep duration (C and D) with incident major adverse cardiovascular events (MACE), with sequential adjustments for potential mediators. |
| Page 19 | Figure S4. Independent associations of step count (A and B) and sleep duration (C and D) with incident major adverse cardiovascular events (MACE), excluding four to six years of follow-up. |
| Page 20 | Figure S5. Independent associations of step count (A and B) and sleep duration (C and D) with incident major adverse cardiovascular events (MACE), excluding prevalent cancer and CVD. |
| Page 21 | Figure S6. Independent associations of step count (A and B) and sleep duration (C and D) with incident major adverse cardiovascular events (MACE), excluding participants with factors that disrupt sleep. |
| Pages 22-23 | Figure S7. Independent associations of step count (A and B) and sleep duration (C and D) with incident major adverse cardiovascular events (MACE), with sequential adjustments for sleep efficiency and step cadence. |
| Page 24 | Figure S8. Joint associations of step count and sleep duration with incident major adverse cardiovascular events (MACE), with additional adjustments for BMI, HbA1c, blood pressure, and total cholesterol. |
| Page 25 | Figure S9. Joint associations of step count and sleep duration with incident major adverse cardiovascular events (MACE), excluding prevalent cancer and CVD. |
| Page 26 | Figure S10. Joint associations of step count and sleep duration with incident major adverse cardiovascular events (MACE), excluding four years of follow-up. |
| Page 27 | Figure S11. Joint associations of step count and sleep duration with incident major adverse cardiovascular events (MACE), with additional adjustments for sleep efficiency and step cadence. |
| Page 28 | Figure S12. Joint associations of step count and sleep duration with incident major adverse cardiovascular events (MACE), excluding participants with factors that disrupt sleep. |

Page 29 References

**Table S1. Search terms and results from literature review.**

| Ref. | Question objective | Key words | Search terms (summary) | MEDLINE | | EMBASE | | |
|---|---|---|---|---|---|---|---|---|
| | | | | Search terms (detailed) | Results | Search terms (detailed) | Results | Total |
| 1 | Exposure | Physical activity and sleep | [Exercise OR Physical Activity OR Steps] AND Sleep | (exp Exercise/ or (physical activity or step* OR exerci*).ti.) and (exp Sleep/ or sleep*.ti.) | 5,016 | (exp Exercise/ or (physical activity or step* OR exerci*).ti.) and (exp Sleep/ or sleep*.ti.) | 15,187 | 20,203 |
| 2 | Outcome | Major cardiovascular events | Cardiovascular disease OR Coronary artery disease OR Cerebrovascular disease OR Peripheral artery disease OR Revascularisation | exp Cardiovascular disease/ or (cardio* or cardia* or heart* or coronary* or myocard* or isch?em*).mp. or exp cerebrovascular disease/ or (stroke* or cerebrovasc* or cerebral vascular).mp. or (peripheral arter* disease* or revasculari* or angioplasty).mp. | 4,259,829 | exp Cardiovascular disease/ or (cardio* or cardia* or heart* or coronary* or myocard* or isch?em*).mp. or exp cerebrovascular disease/ or (stroke* or cerebrovasc* or cerebral vascular).mp. or (peripheral arter* disease* or revasculari* or angioplasty).mp. | 6,751,129 | 11,010,958 |
| 3 | Study design | Observational studies | Case-control OR Retrospective OR Cohort OR Longitudinal OR Prospective OR Cross-sectional OR Prevalence | Case-Control Studies/ or Matched-Pair Analysis/ or (case adj2 control).ti,ab,kf,kw. or cohort studies/ or longitudinal studies/ or follow-up studies/ or prospective studies/ or retrospective studies/ or (cohort or longitudinal or prospective).ti,ab,kf,kw. or Cross-Sectional Studies/ or Prevalence/ or (association adj2 (studies or study)).mp. or (ecological adj (studies or study)).mp. or (cross-sectional or prevalence or transversal).ti,ab,kf,kw. or (association or associations).ti. | 5,017,406 | Case-Control Studies/ or Matched-Pair Analysis/ or (case adj2 control).ti,ab,kf,kw. or cohort studies/ or longitudinal studies/ or follow-up studies/ or prospective studies/ or retrospective studies/ or (cohort or longitudinal or prospective).ti,ab,kf,kw. or Cross-Sectional Studies/ or Prevalence/ or (association adj2 (studies or study)).mp. or (ecological adj (studies or study)).mp. or (cross-sectional or prevalence or transversal).ti,ab,kf,kw. or (association or associations).ti. | 7,157,697 | 12,175,103 |
| 4 | Population | Adult humans | Humans NOT child/infant | NOT (exp child/ or exp infant/) AND limit to humans | NA | NOT (exp child/ or exp infant/) AND limit to humans | NA | NA |
| 5 | Additional criteria | English language only Excluding conference abstracts | English language Conference abstracts | NOT conference abstract.pt and limit to english language | NA | NOT conference abstract.pt and limit to English language | NA | NA |
| **Combined** | | | | **1 AND 2 AND 3 AND 4 AND 5** | 287 | **1 AND 2 AND 3 AND 4 AND 5** | 1219 | 1506 |

**Table S2. Summary of study characteristics for articles included in literature review.**

| Study authors (publication year) | Study name (country) | Mean/median age | Sample size | % Female | Event number | Mean/median follow-up years | Ascertainment of physical activity and sleep |
|---|---|---|---|---|---|---|---|
| **Cohort studies** | | | | | | | |
| Liang et al. (2023)[1] | UK Biobank (United Kingdom) | 62·4 | 92,221 | 56·4% | 1,074 | 7·0* | Wrist-worn Axivity AX3 accelerometer for 7 days |
| Huang et al. (2022)[2] | UK Biobank (United Kingdom) | 55·9 | 380,055 | 55·9% | 4,095 | 11·1 | Self-report questionnaire |
| Chen et al. (2022)[3] | MJ Cohort (Taiwan) | 39·7 | 341,248 | 51·7% | 3,845 | 15·0 | Self-report questionnaire |
| Bellavia et al. (2013)[4] | Cohort of Swedish Men and Swedish Mammography Cohort (Sweden) | 60·1 | 70,973 | 47·0% | 3,981 | 15·0 | Self-report questionnaire |
| Wang et al. (2023)[5] | National Health and Nutrition Examination Survey (United States) | 49·0* | 29,058 | 51·5% | 1,001 | 7·4 | Self-report questionnaire |
| Wennman et al. (2017)[6] | Finnish Former Elite Athletes' Cohort and military conscription register (Finland) | 55·0 | 1,638 | 0% | 391 | 26·0 | Self-report questionnaire |
| Xiao et al. (2014)[7] | National Institutes of Health-AARP Diet and Health Study (United States) | 62·0 | 239,896 | 44% | 11,635 | 14·0 | Self-report questionnaire |
| **Cross-sectional studies** | | | | | | | |
| Huang et al. (2021)[8] | Tianning Cohort (China) | 51·0 | 5,130 | 58·7% | 531 | NA | Self-report questionnaire |

* The values shown represent the median.

**Table S3. Summary of main results for articles included in literature review.**

| Study authors (publication year) | Classification of PA | Classification of sleep | Outcomes | RR/OR/HR | | | | Interaction | Adjustments |
|---|---|---|---|---|---|---|---|---|---|
| **Cohort studies** | | | | | | | | | |
| Liang et al. (2023)[1] | By weekly average vector magnitude:<br><br>Low PA: ≤24·0 m*g*<br><br>Medium PA: 24·0-30·4 m*g*<br><br>High PA: 30·4+ m*g* | By daily sleep duration:<br><br>Short sleep: <6 h<br><br>Normal sleep: 6-8 h<br><br>Long sleep: 8+ h | Death from CVD<br><br>ICD-10: I00-99 | | **Low PA**<br>Short sleep:<br>HR 3·93<br>(2·90-5·32)<br><br>Normal sleep:<br>HR 2·27<br>(1·82-2·84)<br><br>Long sleep:<br>HR 2·35<br>(1·84-3·01) | **Medium PA**<br>Short sleep:<br>HR 1·77<br>(1·16-2·69)<br><br>Normal sleep:<br>HR 1·38<br>(1·08-1·75)<br><br>Long sleep:<br>HR 1·09<br>(0·75-1·59) | **High PA**<br>Short sleep:<br>HR 1·40<br>(0·88-2·25)<br><br>Normal sleep:<br>Ref<br><br>Long sleep:<br>HR 0·49<br>(0·24-1·01) | Significant interaction observed ($p$<0·05) | Age<br>Sex<br>Ethnicity<br>Recruitment centre<br>Education<br>Season accelerometer worn<br>SES<br>BMI<br>Diet-related factors<br>Smoking status<br>Alcohol intake<br>Work shift history |
| Huang et al. (2022)[2] | By weekly MET-mins:<br><br>No MVPA<br><br>Low PA:<br>0 - <600 MET-mins/week<br><br>Medium PA:<br>600 - <1200 MET-mins/week<br><br>High PA:<br>1200+ MET-mins/week | By composite sleep health score (out of 5):<br><br>Poor sleep: ≤1<br><br>Intermediate sleep: 2-3<br><br>Healthy sleep: 4+ | Death from CVD, excl. hypertension and diseases of arteries and lymph | **No MVPA**<br>Poor sleep:<br>HR 1·67<br>(1·27-2·19)<br><br>Intermediate sleep:<br>HR 1·50<br>(1·34-1·67)<br><br>Healthy sleep:<br>HR 1·22<br>(1·07-1·38) | **Low PA**<br>Poor sleep:<br>HR 1·91<br>(1·31-2·78)<br><br>Intermediate sleep:<br>HR 1·11<br>(0·96-1·30)<br><br>Healthy sleep:<br>HR 1·11<br>(0·95-1·30) | **Medium PA**<br>Poor sleep:<br>HR 1·33<br>(0·86-2·05)<br><br>Intermediate sleep:<br>HR 1·14<br>(1·00-1·30)<br><br>Healthy sleep:<br>HR 1·06<br>(0·93-1·21) | **High PA**<br>Poor sleep:<br>HR 1·37<br>(1·09-1·72)<br><br>Intermediate sleep:<br>HR 1·07<br>(0·98-1·16)<br><br>Healthy sleep:<br>Ref | Significant interaction observed ($p$<0·001) | Age<br>Sex<br>BMI<br>SES<br>Fruit and vegetable intake<br>Sedentary behaviour<br>Mental health issues<br>Smoking status<br>Employment status<br>Alcohol intake |
| Chen et al. (2022)[3] | By weekly MET-hrs:<br><br>Low PA:<br><7·5 MET-h/week<br><br>Medium PA:<br>7·5-14·9 MET-h/week<br><br>High PA:<br>15-29·9 MET-h/week | By daily sleep duration:<br><br>Short sleep: <6 h<br><br>Normal sleep: 6-8 h<br><br>Long sleep: 8+ h | Death from CVD<br><br>ICD-10: I01-I02, I05-I09, I10-I15, I20-I25, I27, I30-I52, I60-I69, I70, I71 | | **Low PA**<br>Short sleep:<br>HR 1·26<br>(1·10-1·45)<br><br>Normal sleep:<br>HR 1·22<br>(1·08-1·38)<br><br>Long sleep:<br>HR 1·66<br>(1·43-1·92) | **Medium PA**<br>Short sleep:<br>HR 1·14<br>(0·99-1·32)<br><br>Normal sleep:<br>HR 1·15<br>(0·96-1·38)<br><br>Long sleep:<br>HR 1·29<br>(1·02-1·63) | **High PA**<br>Short sleep:<br>HR 1·04<br>(0·87-1·24)<br><br>Normal sleep:<br>Ref<br><br>Long sleep:<br>HR 1·02<br>(0·81-1·29) | Significant interaction observed (no $p$-value reported) | Age<br>Sex<br>Education<br>Marital status<br>Smoking status<br>Alcohol intake<br>Fruit and vegetable intake<br>Physical health status |

| | | | | | | | | | |
|---|---|---|---|---|---|---|---|---|---|
| Bellavia et al. (2013)[4] | By daily MET-hrs (in tertiles):<br><br>Low PA:<br><39·3 MET-h/day<br><br>Medium PA:<br>39·3-44·2 MET-h/day<br><br>High PA:<br>44·2+ MET-h/day | By daily sleep duration:<br><br><6 h<br><br>6-6·5 h<br><br>6·6-7·4 h<br><br>7·5-8 h<br><br>8+ h | Death from CVD<br><br>Did not specify diagnosis codes. | | **Low PA**<br><6 h:<br>HR 1·45<br>(0·85-2·22)<br><br>6-6·5 h:<br>HR 1·54<br>(1·25-1·90)<br><br>6·6-7·4 h:<br><u>Ref</u><br><br>7·5-8 h:<br>HR 1·17<br>(0·99-1·37)<br><br>8+ h:<br>HR 1·41<br>(1·14-1·74) | **Medium PA**<br><6 h:<br>HR 1·23<br>(0·89-1·70)<br><br>6-6·5 h:<br>HR 1·05<br>(0·85-1·28)<br><br>6·6-7·4 h:<br><u>Ref</u><br><br>7·5-8 h:<br>HR 0·91<br>(0·76-1·09)<br><br>8+ h:<br>HR 0·84<br>(0·60-1·17) | **High PA**<br><6 h:<br>HR 1·54<br>(1·19-2·01)<br><br>6-6·5 h:<br>HR 1·15<br>(0·95-1·40)<br><br>6·6-7·4 h:<br><u>Ref</u><br><br>7·5-8 h<br>HR 0·97<br>(0·80-1·17)<br><br>8+ h:<br>HR 0·73<br>(0·43-1·23) | Significant interaction observed ($p$<0·001) | Age<br>Sex<br>BMI<br>Smoking status<br>Alcohol intake<br>Education |
| Wang et al. (2023)[5] | By weekly MET-mins:<br><br>Low PA:<br>0 - <600 MET-mins/week<br><br>Medium PA:<br>600 - <1200 MET-mins/week<br><br>High PA:<br>1200+ MET-mins/week | By daily sleep duration:<br><br><5·5 h<br><br>5·5 - <6·5 h<br><br>6·5 - <7·5 h<br><br>7·5 - <8·5 h<br><br>8·5+ h | Death from CVD<br><br>ICD-10:<br>I00-I09, I11, I13, I20-I51, I60-I69 | | **Low PA**<br><5·5 h:<br>HR 3·35<br>(2·30-4·90)<br><br>5·5 - <6·5 h:<br>HR 2·71<br>(1·86-3·94)<br><br>6·5 - <7·5 h:<br>HR 2·43<br>(1·64-3·59)<br><br>7·5 - <8·5 h:<br>HR 2·49<br>(1·79-3·44)<br><br>8·5+ h:<br>HR 3·08<br>(2·12-4·46) | **Medium PA**<br><5·5 h:<br>HR 0·87<br>(0·48-1·55)<br><br>5·5 - <6·5 h:<br>HR 1·71<br>(1·09-2·69)<br><br>6·5 - <7·5 h:<br>HR 2·63<br>(1·45-4·76)<br><br>7·5 - <8·5 h:<br>HR 2·02<br>(1·08-3·77)<br><br>8·5+ h:<br>HR 1·41<br>(1·14-1·74) | **High PA**<br><5·5 h:<br>HR 3·03<br>(1·77-5·16)<br><br>5·5 - <6·5 h:<br>HR 1·80<br>(1·15-2·82)<br><br>6·5 - <7·5 h:<br><u>Ref</u><br><br>7·5 - <8·5 h:<br>HR 1·92<br>(1·10-3·35)<br><br>8·5+ h:<br>HR 1·86<br>(1·01-3·4) | No significant interaction observed ($p$=0.173) | Age<br>Sex<br>BMI<br>Education<br>Marital status<br>Employment status<br>Smoking status<br>Alcohol intake<br>Depression<br>Sedentary behaviour<br>Baseline chronic diseases |
| Wennman et al. (2017)[6] | By weekly leisure MET-mins:<br><br>Insufficient PA:<br>(<450 MET-mins/week)<br><br>Sufficient PA: (450+ MET-mins/week) | By daily sleep duration:<br><br>Short sleep:<br><6 h<br><br>Mid-range sleep:<br>6·5-8·5 h | Death from CVD<br><br>Did not specify diagnosis codes. | | | **Insufficient PA**<br>Short sleep:<br>HR 1·98<br>(1·25-3·12)<br><br>Mid-range sleep:<br>HR 1·18 | **Sufficient PA**<br><br>Short sleep:<br>HR 0·75<br>(0·44-1·30)<br><br>Mid-range sleep:<br><u>Ref</u> | Significant interaction observed ($p$=0.01) | Occupation<br>Marital status<br>Smoking status<br>Alcohol intake<br>BMI<br>Life satisfaction<br>Chronotype<br>Sleep medication<br>Chronic diseases |

| | | | | | | | | | |
|---|---|---|---|---|---|---|---|---|---|
| | | Long sleep:<br>9+ h | | | | (0·92-1·52)<br><br>Long sleep:<br>HR 0·75<br>(0·47-1·18) | Long sleep:<br>HR 0·96<br>(0·65-1·42) | | |
| Xiao et al. (2014)[7] | By weekly MVPA time:<br><br>Low PA:<br><1 h/week<br><br>Medium PA:<br>1-3 h/week<br><br>High PA:<br>4+ h/week | By daily sleep duration:<br><br><5 h<br><br>5-6·9 h<br><br>7-8·9 h<br><br>9+ h | Death from CVD<br><br>ICD-10:<br>I00-I78 | | **Low PA**<br><5 h:<br>HR 1·28<br>(1·17-1·40)<br><br>5-6·9 h:<br>HR 1·02<br>(0·99-1·06)<br><br>7-8·9 h:<br>Ref<br><br>9+ h:<br>HR 1·20<br>(1·10-1·30) | **Medium PA**<br><5 h:<br>HR 1·23<br>(1·10-1·38)<br><br>5-6·9 h:<br>HR 1·07<br>(1·02-1·11)<br><br>7-8·9 h:<br>Ref<br><br>9+ h:<br>HR 1·07<br>(0·96-1·18) | **High PA**<br><5 h:<br>HR 1·13<br>(1·04-1·23)<br><br>5-6·9 h:<br>HR 1·06<br>(1·02-1·09)<br><br>7-8·9 h:<br>Ref<br><br>9+ h:<br>HR 1·10<br>(1·02-1·19) | No significant interaction observed<br>(*p*=0.51) | Age<br>Sex<br>Ethnicity<br>Marital status<br>Education<br>Self-reported health<br>Smoking status<br>Smoking dose<br>Years since quitting smoking<br>Alcohol intake |
| **Cross-sectional studies** | | | | | | | | | |
| Huang et al. (2021)[8] | By weekly MET-mins:<br><br>Inactive:<br><600 MET-mins/week<br><br>Active:<br>600+ MET-mins/week | By daily sleep duration:<br><br><9 h<br><br>9+ h | Peripheral artery disease from ABPI measurement of <0·9 | | | **Inactive**<br><9 h:<br>OR 1·29<br>(1·04-1·60)<br><br>9+ h:<br>OR 2·41<br>(1·55-3·76) | **Active**<br><9 h:<br>Ref<br><br>9+ h:<br>OR 1·16<br>(0·79-1·71) | Significant interaction observed<br>(no *p*-value reported) | Age<br>Sex<br>Education<br>Depressive symptoms<br>Obstructive sleep apnoea<br>Cigarette smoking<br>Alcohol intake<br>BMI<br>SBP<br>DBP<br>Diabetes mellitus<br>Total cholesterol<br>LDL cholesterol |

PA, physical activity; RR, relative risk; OR, odds ratio; HR, hazard ratio; MG, milli-gravity unit; CVD, cardiovascular disease; SES, socioeconomic status; BMI, body mass index; MET, Metabolic Equivalent of Task; MVPA, moderate-vigorous physical activity; ABPI, Ankle Brachial Pressure Index; SBP, systolic blood pressure; DBP, diastolic blood pressure; LDL, low-density lipoprotein.

**Table S4. Medical classification codes for incident major adverse cardiovascular events (MACE; ICD-10), previous disease (ICD-10), and revascularisation procedures (OPCS-4).**

| Disease category | Description | ICD-10 code | ICD-9 code | OPCS-4 code |
|---|---|---|---|---|
| **Outcomes** | | | | |
| Myocardial infarction | Acute myocardial infarction | I21 | 410 | |
| | Subsequent myocardial infarction | I22 | 411 | |
| Stroke | Subarachnoid haemorrhage | I60 | 430 | |
| | Intracerebral haemorrhage | I61 | 431 | |
| | Other nontraumatic intracranial haemorrhage | I62 | 432 | |
| | Cerebral infarction | I63 | 433-434 | |
| | Stroke, not specified as haemorrhage or infarction | I64 | 436 | |
| Cardiovascular death | Death from diseases of the circulatory system | I00-I99 | 390-459 | |
| **Exclusions** | | | | |
| Cancer | Malignant neoplasms | C00-C43, C45-C97 | 130-172, 174-239 | |
| Cardiovascular disease | Diseases of the circulatory system | I00-I99 | 390-459 | |
| Other specified extrapyramidal and movement disorders | Restless legs syndrome | G25.8 | 333.99 | |
| Sleep apnoea | Sleep apnoea | G47.3 | 327.2 | |
| **Revascularisation procedures** | | | | |
| Procedures of the heart | Revascularisation of wall of heart | | | K23.4 |
| | Coronary artery revascularisation procedures (excluding repair of coronary artery, other open operations on coronary artery, transluminal balloon angioplasty of coronary artery, and diagnostic transluminal operations on coronary artery) | | | K40-K46, K49-K50 |
| | Percutaneous transluminal balloon angioplasty and insertion of stent into coronary artery | | | K75 |
| Procedures of the arteries and veins | Aorta revascularisation procedures (excluding aortography) | | | L16, L18-L23, L25, L26.0-L26.3, L26.5-L26.9, L27-L28 |
| | Carotid and subclavian artery revascularisation procedures (excluding cerebral artery procedures, arteriography of carotid artery, percutaneous transluminal embolization of subclavian artery, and arteriography of subclavian artery) | | | L29-L30, L31.0-L31.1, L31.3-L31.9, L37-L38, L39.0-L39.2, L39.5-L39.9 |
| | Iliac and femoral artery procedures (excluding arteriography of iliac artery, percutaneous transluminal embolization of femoral artery, and arteriography of femoral artery) | | | L48-L53, L54.0-L54.2, L54.4-L54.9, L56-L62, L63.0-L63.2, L63.5-L63.9 |
| | Other therapeutic transluminal operations on artery | | | L66 |
| | Repair of other artery | | | L68 |

ICD, International Classification of Diseases; OPCS, Office of Population Censuses and Surveys Classification of Interventions and Procedures.

**Table S5. UK Biobank fields for all variables used.**

| Variable | UK Biobank field | Source | Time of collection | Additional information |
|---|---|---|---|---|
| **Exposures** | | | | |
| Accelerometer-measured step count and sleep duration | 90001 | Accelerometer data | Entry to accelerometer study | Derived from raw data as described in the Methods |
| **Covariates** | | | | |
| Age at end of accelerometer wear | 34, 90001 | Baseline characteristics, accelerometer data | Initial entry into UK Biobank, entry to accelerometer study | Estimated from the date at end of accelerometer wear time and date of birth (with the 15th imputed as the day of birth for all participants) |
| Sex | 31 | Baseline characteristics | Initial entry into UK Biobank | |
| Ethnicity | 21000 | Baseline characteristics | Initial entry into UK Biobank | |
| Townsend Deprivation Index | 22189 | Baseline characteristics | Initial entry into UK Biobank | |
| Education | 6138 | Baseline characteristics | Initial entry into UK Biobank | |
| Smoking status | 20116 | Baseline characteristics | Initial entry into UK Biobank | |
| Alcohol intake | 1558 | Baseline characteristics | Initial entry into UK Biobank | |
| Processed meat intake | 1349 | Baseline characteristics | Initial entry into UK Biobank | |
| Body mass index | 21001 | Physical measures | Initial entry into UK Biobank | Constructed from height and weight measurements taken from the participant |
| Glycated haemoglobin | 30750 | Blood biochemistry | Initial entry into UK Biobank | |
| Systolic and diastolic blood pressure | 94, 4079, 4080, 93 | Physical measures | Initial entry into UK Biobank | |
| Total cholesterol | 23400 | Nuclear magnetic resonance metabolomics | Initial entry into UK Biobank | |
| Accelerometer-measured step cadence | 90001 | Accelerometer data | Entry to accelerometer study | Derived from raw data as described in the Methods |
| Accelerometer-measured sleep efficiency | 90001 | Accelerometer data | Entry to accelerometer study | Derived from raw data as described in the Methods |
| **Outcome** | | | | |
| Date of first incident major adverse cardiovascular event | 2002, 100093 | Hospital inpatient data, death register | Ongoing follow-up | |
| Date of censoring | 54, 100093, 191 | Death register, ongoing characteristics, UK Biobank dates of data availability | Ongoing follow-up | Specific censoring dates according to assessment centre:<br>England – 31st of October 2022<br>Scotland – 31st of August 202<br>Wales – 31st of May 2022 |
| **Exclusions** | | | | |
| Hospital diagnosis of previous disease | 2002 | Hospital inpatient data | Ongoing follow-up | |
| Self-reported previous disease | 20001, 20002, 6150 | Verbal interview | Initial entry into UK Biobank, first repeat assessment visit, imaging visit | |
| Self-reported medication | 20003 | Verbal interview | Initial entry into UK Biobank | |
| Self-reported shift work | 3426 | Baseline characteristics | Initial entry into UK Biobank | |
| Daylight saving time changes | 90001 | Accelerometer data | Entry to accelerometer study | |

Initial entry into UK Biobank between 2006-2010. Entry to accelerometer study between 2013-2015. First repeat assessment visit between 2012-2013. Imaging visit from 2014 onwards.

**Table S6. Hazard ratios and their respective conventional and floating absolute risk confidence intervals for the joint associations of step count and sleep duration with incident major adverse cardiovascular events (MACE).**

| Participant category by step count and sleep duration | Maximally adjusted model | | | | Additional adjustments for step cadence and sleep efficiency | | | | Additional adjustments for potential mediators | | | |
|---|---|---|---|---|---|---|---|---|---|---|---|---|
| | **No. of events** | **HR** | **Conventional 95% CI** | **FAR 95% CI** | **No. of events** | **HR** | **Conventional 95% CI** | **FAR 95% CI** | **No. of events** | **HR** | **Conventional 95% CI** | **FAR 95% CI** |
| **Low step count** | | | | | | | | | | | | |
| Short sleep duration | 580 | 1·84 | 1·62-2·10 | 1·70-2·00 | 580 | 1·53 | 1·32-1·78 | 1·40-1·68 | 498 | 1·63 | 1·41-1·87 | 1·49-1·78 |
| Medium sleep duration | 536 | 1·38 | 1·21-1·58 | 1·27-1·50 | 536 | 1·16 | 0·99-1·34 | 1·05-1·27 | 479 | 1·29 | 1·12-1·48 | 1·18-1·42 |
| Long sleep duration | 425 | 1·56 | 1·34-1·79 | 1·41-1·71 | 425 | 1·31 | 1·12-1·52 | 1·18-1·45 | 377 | 1·47 | 1·26-1·70 | 1·33-1·63 |
| **Medium step count** | | | | | | | | | | | | |
| Short sleep duration | 496 | 1·47 | 1·28-1·68 | 1·34-1·60 | 496 | 1·36 | 1·19-1·57 | 1·25-1·49 | 447 | 1·41 | 1·21-1·60 | 1.28-1.54 |
| Medium sleep duration | 574 | 1·21 | 1·06-1·38 | 1·11-1·31 | 574 | 1·13 | 0·99-1·29 | 1·04-1·23 | 503 | 1·16 | 1·02-1·34 | 1.06-1.27 |
| Long sleep duration | 312 | 1·19 | 1·03-1·39 | 1·07-1·33 | 312 | 1·12 | 0·96-1·30 | 1·00-1·25 | 285 | 1·20 | 1·02-1·40 | 1.07-1.35 |
| **High step count** | | | | | | | | | | | | |
| Short sleep duration | 356 | 1·15 | 0·99-1·33 | 1·03-1·27 | 356 | 1·14 | 0·98-1·32 | 1·02-1·27 | 319 | 1·13 | 0·98-1·33 | 1·01-1·26 |
| Medium sleep duration | 375 | 1·00 | Reference | 0·90-1·11 | 375 | 1·00 | Reference | 0·90-1·12 | 332 | 1·00 | Reference | 0·90-1·11 |
| Long sleep duration | 163 | 0·97 | 0·81-1·17 | 0·83-1·13 | 163 | 0·97 | 0·81-1·17 | 0·83-1·14 | 137 | 0·92 | 0·75-1·12 | 0·78-1·09 |
| **$P_{interaction}$** | 0·42 | | | | 0·44 | | | | 0·54 | | | |
| | **Excluding first 4 years of follow-up** | | | | **Excluding all prevalent cancer and CVD (incl. self-reported)** | | | | **Excluding participants with factors that disrupt sleep** | | | |
| | **No. of events** | **HR** | **Conventional 95% CI** | **FAR 95% CI** | **No. of events** | **HR** | **Conventional 95% CI** | **FAR 95% CI** | **No. of events** | **HR** | **Conventional 95% CI** | **FAR 95% CI** |
| **Low step count** | | | | | | | | | | | | |
| Short sleep duration | 325 | 1·67 | 1·41-1·99 | 1·50-1·87 | 171 | 1·41 | 1·15-1·73 | 1·21-1·64 | 531 | 1·87 | 1·63-2·15 | 1·72-2·04 |
| Medium sleep duration | 294 | 1·21 | 1·02-1·44 | 1·08-1·36 | 182 | 1·10 | 0·90-1·34 | 0·95-1·27 | 486 | 1·36 | 1·19-1·56 | 1·25-1·49 |
| Long sleep duration | 230 | 1·32 | 1·10-1·59 | 1·16-1·50 | 142 | 1·28 | 1·03-1·59 | 1·09-1·51 | 387 | 1·53 | 1·32-1·77 | 1·39-1·69 |
| **Medium step count** | | | | | | | | | | | | |
| Short sleep duration | 271 | 1·34 | 1·12-1·19 | 1·19-1·51 | 207 | 1·30 | 1·07-1·58 | 1·13-1·49 | 454 | 1·48 | 1·28-1·70 | 1·35-1·62 |
| Medium sleep duration | 332 | 1·15 | 0·97-1·36 | 1·03-1·28 | 263 | 1·12 | 0·93-1·35 | 0·99-1·27 | 538 | 1·22 | 1·07-1·40 | 1·12-1·33 |
| Long sleep duration | 182 | 1·12 | 0·92-1·36 | 0·96-1·29 | 139 | 1·10 | 0·88-1·36 | 0·93-1·30 | 288 | 1·18 | 1·00-1·38 | 1·05-1·32 |
| **High step count** | | | | | | | | | | | | |
| Short sleep duration | 207 | 1·15 | 0·95-1·40 | 1·01-1·32 | 137 | 0·86 | 0·69-1·06 | 0·72-1·01 | 329 | 1·16 | 0·99-1·34 | 1·04-1·29 |
| Medium sleep duration | 221 | 1·00 | Reference | 0·88-1·14 | 203 | 1·00 | Reference | 0·87-1·15 | 350 | 1·00 | Reference | 0·90-1·11 |
| Long sleep duration | 88 | 0·87 | 0·68-1·12 | 0·71-1·08 | 75 | 0·87 | 0·67-1·14 | 0·70-1·10 | 147 | 0·93 | 0·77-1·13 | 0·79-1·10 |
| **$P_{interaction}$** | 0·44 | | | | 0·13 | | | | 0·31 | | | |

Participants were stratified into nine mutually exclusive groups using the following classifications for median daily step count (low [<7500], intermediate [7500-11000], high [11000+; reference) and median overnight sleep duration (short [<6.5 hours], intermediate [6.5-7.5 hours; reference], long [7.5+ hours]).

The maximally adjusted model adjusted for age, sex, ethnicity, education, TDI, smoking status, alcohol intake, and processed meat intake.

Potential mediators adjusted for included BMI, HbA1c, blood pressure, and total cholesterol.

Factors considered to disrupt sleep included self-reported shift work, a prior diagnosis of sleep apnoea or restless leg syndrome, and accelerometer wear that overlapped with daylight saving time changes.

The *p*-value for interaction was derived from a likelihood ratio test comparing the maximally adjusted model with and without an interaction term between step count and sleep duration.

HR, hazard ratio; 95% CI, 95% confidence interval; FAR, floating absolute risk; TDI, Townsend Deprivation Index; BMI, body mass index; HbA1c, glycated haemoglobin.

**Table S7. STROBE statement checklist.**

| | Item number | Recommendation | Page number |
|---|---|---|---|
| **Title and abstract** | 1 | (*a*) Indicate the study's design with a commonly used term in the title or the abstract | 1, 2 |
| | | (*b*) Provide in the abstract an informative and balanced summary of what was done and what was found | 2 |
| **Introduction** | | | |
| Background/rationale | 2 | Explain the scientific background and rationale for the investigation being reported | 6<br>Tables S1-S3 |
| Objectives | 3 | State specific objectives, including any prespecified hypotheses | 6 |
| **Methods** | | | |
| Study design | 4 | Present key elements of study design early in the paper | 7 |
| Setting | 5 | Describe the setting, locations, and relevant dates, including periods of recruitment, exposure, follow-up, and data collection | 7, 8<br>Table S5 |
| Participants | 6 | (*a*) Give the eligibility criteria, and the sources and methods of selection of participants. Describe methods of follow-up | 7, 8<br>Table S5 |
| | | (*b*) For matched studies, give matching criteria and number of exposed and unexposed | NA |
| Variables | 7 | Clearly define all outcomes, exposures, predictors, potential confounders, and effect modifiers. Give diagnostic criteria, if applicable | 7-9<br>Tables S4-S5 |
| Data sources/measurement | 8* | For each variable of interest, give sources of data and details of methods of assessment (measurement). Describe comparability of assessment methods if there is more than one group | 7-9<br>Table S5 |
| Bias | 9 | Describe any efforts to address potential sources of bias | 8, 9, 10-11 |
| Study size | 10 | Explain how the study size was arrived at | 7 |
| Quantitative variables | 11 | Explain how quantitative variables were handled in the analyses. If applicable, describe which groupings were chosen and why | 9 |
| Statistical methods | 12 | (*a*) Describe all statistical methods, including those used to control for confounding | 9-10 |
| | | (*b*) Describe any methods used to examine subgroups and interactions | 10 |
| | | (*c*) Explain how missing data were addressed | 7, 11 |
| | | (*d*) If applicable, explain how loss to follow-up was addressed | 11 |
| | | (*e*) Describe any sensitivity analyses | 10-11 |
| **Results** | | | |
| Participants | 13* | (*a*) Report numbers of individuals at each stage of study – e.g., numbers potentially eligible, examined for eligibility, confirmed eligible, included in the study, completing follow-up, and analysed | 11<br>Figure 1 |
| | | (*b*) Give reasons for non-participation at each stage | 11<br>Figure 1 |
| | | (*c*) Consider use of a flow diagram | Figure 1 |
| Descriptive data | 14* | (*a*) Give characteristics of study participants (e.g., demographic, clinical, social) and information on exposures and potential confounders | 11-12<br>Table 1 |
| | | (*b*) Indicate number of participants with missing data for each variable of interest | Table 1 |
| | | (*c*) Summarise follow-up time (e.g., average and total amount) | 12 |
| Outcome data | 15* | Report numbers of outcome events or summary measures over time | 12 |
| Main results | 16 | (*a*) Give unadjusted estimates and, if applicable, confounder-adjusted estimates and their precision (e.g., 95% confidence interval). Make clear which confounders were adjusted for and why they were included | Figure 2<br>Figure 3<br>Figure S1 |
| | | (*b*) Report category boundaries when continuous variables were categorised | Figure 2<br>Figure 3 |
| | | (*c*) If relevant, consider translating estimates of relative risk into absolute risk for a meaningful time period | Figure 2<br>Figure 3 |
| Other analyses | 17 | Report other analyses done—e.g., analyses of subgroups and interactions, and sensitivity analyses | Table S6<br>Figures S2-S12 |
| **Discussion** | | | |
| Key results | 18 | Summarise key results with reference to study objectives | 14, 17 |
| Limitations | 19 | Discuss limitations of the study, taking into account sources of potential bias or imprecision. Discuss both direction and magnitude of any potential bias | 17 |
| Interpretation | 20 | Give a cautious overall interpretation of results considering objectives, limitations, multiplicity of analyses, results from similar studies, and other relevant evidence | 14-17 |
| Generalisability | 21 | Discuss the generalisability (external validity) of the study results | 17 |
| **Other information** | | | |

| | | | |
|---|---|---|---|
| Funding | 22 | Give the source of funding and the role of the funders for the present study and, if applicable, for the original study on which the present article is based | 3, 11 |

* Information given separately for exposed and unexposed groups.

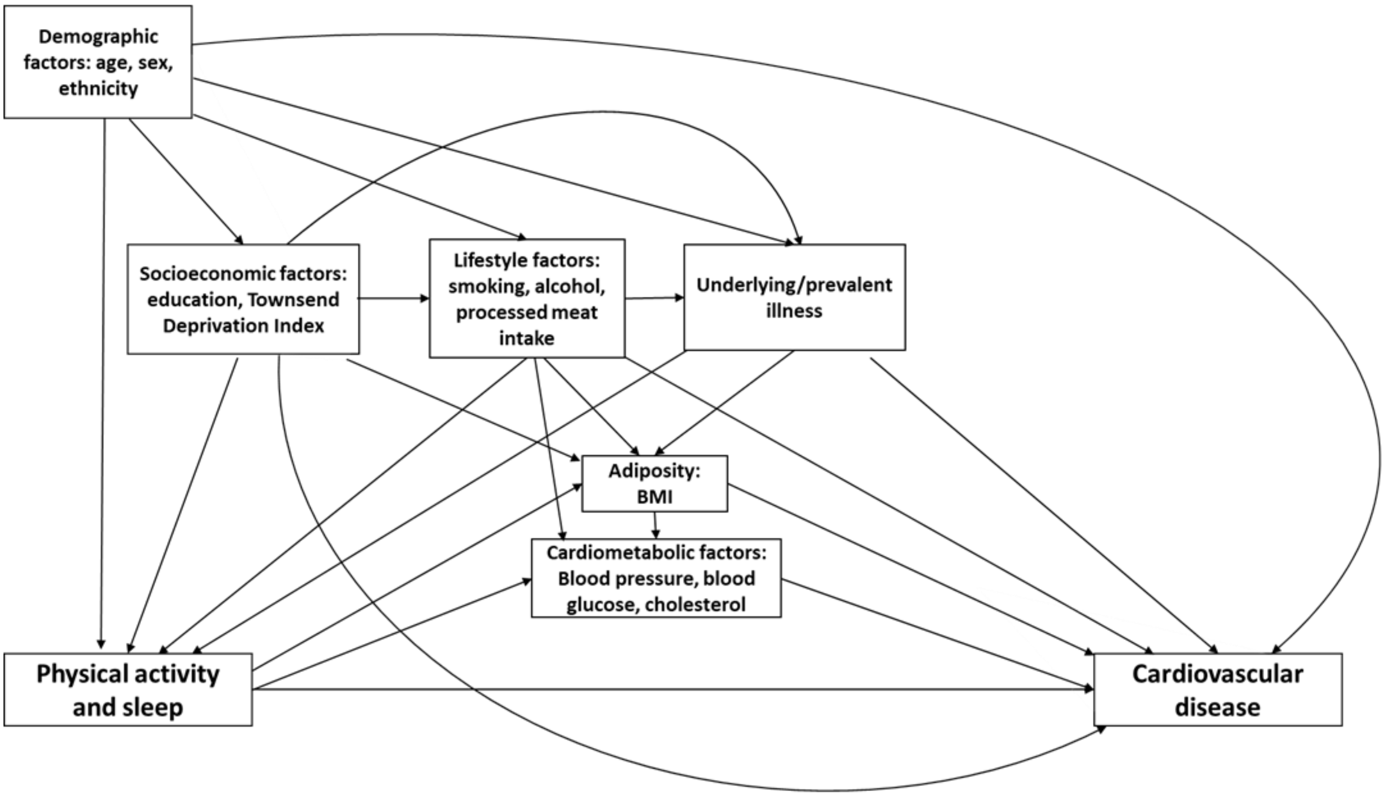


**Figure S1. Causal diagram depicting the mechanistic pathways between physical activity, sleep, key covariates, and cardiovascular disease.**

Covariates identified with a potential confounding effect were age, sex, ethnicity, education, TDI, smoking status, alcohol intake, and processed meat intake.

Covariates identified with a potential mediating effect were BMI, blood pressure, HbA1c, and total cholesterol.

Additional potential confounding effects of underlying and prevalent illness were examined by exclusions from the study population.

TDI, Townsend Deprivation Index; BMI, body mass index; HbA1c, glycated haemoglobin.

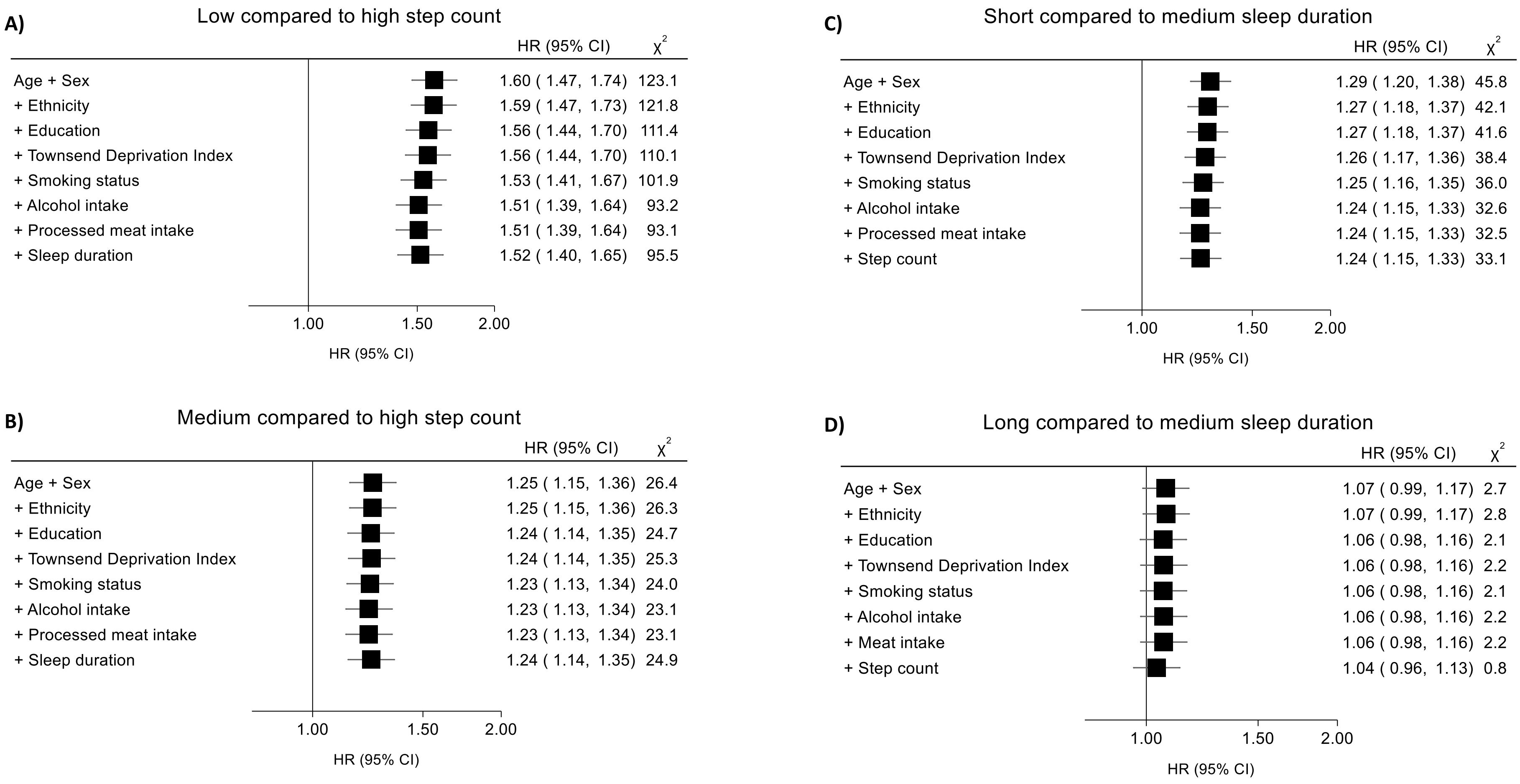


**Figure S2. Independent associations of step count (A and B) and sleep duration (C and D) with incident major adverse cardiovascular events (MACE), with sequential adjustments for potential confounders.**

HRs after sequential adjustments for potential confounders have been depicted for each participant group according to the following classifications for median daily step count (low [<7500], intermediate [7500-11000], high [11000+; reference) and median overnight sleep duration (short [<6.5 hours], intermediate [6.5-7.5 hours; reference], long [7.5+ hours]). $X^2$ values were derived from likelihood ratio tests comparing models with and without each covariate, and represent the potential confounding effect of that covariate.

The horizontal lines represent 95% CIs based on floating absolute risks. The size of each box is relative to the amount of statistical information available. The HRs for the x-axis have been plotted on a log scale.

HR, hazard ratio; 95% CI, 95% confidence interval.

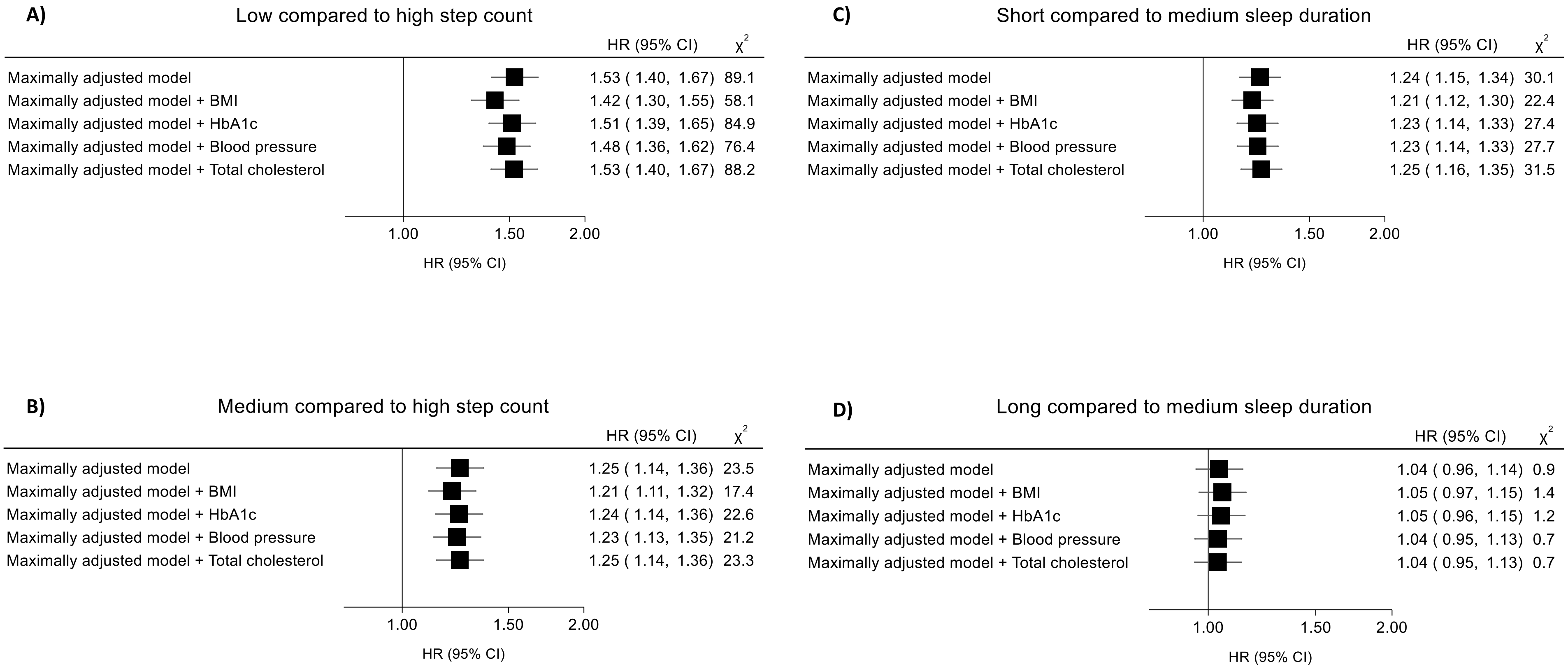


**Figure S3. Independent associations of step count (A and B) and sleep duration (C and D) with incident major adverse cardiovascular events (MACE), with sequential adjustments for potential mediators.**

Additional participants were excluded due to missing covariate data, resulting in a study subpopulation of 78,866 individuals.

HRs after sequential adjustments for potential mediators have been depicted for each participant group according to the following classifications for median daily step count (low [<7500], intermediate [7500-11000], high [11000+; reference) and median overnight sleep duration (short [<6.5 hours], intermediate [6.5-7.5 hours; reference], long [7.5+ hours]). $X^2$ values were derived from likelihood ratio tests comparing the maximally adjusted model with and without each covariate, and represent the potential confounding effect of that covariate. The maximally adjusted model adjusted for age, sex, ethnicity, education, TDI, smoking status, alcohol intake, processed meat intake, and sleep duration or step count.

For participants who self-reported taking medication for diabetes mellitus, hypertension, or hypercholesterolaemia, biomarker values were adjusted based on mean treatment effects reported in the literature.[9-11]

The horizontal lines represent 95% CIs based on floating absolute risks. The size of each box is relative to the amount of statistical information available. The HRs for the x-axis have been plotted on a log scale.

HR, hazard ratio; TDI, Townsend Deprivation Index; 95% CI, 95% confidence interval.

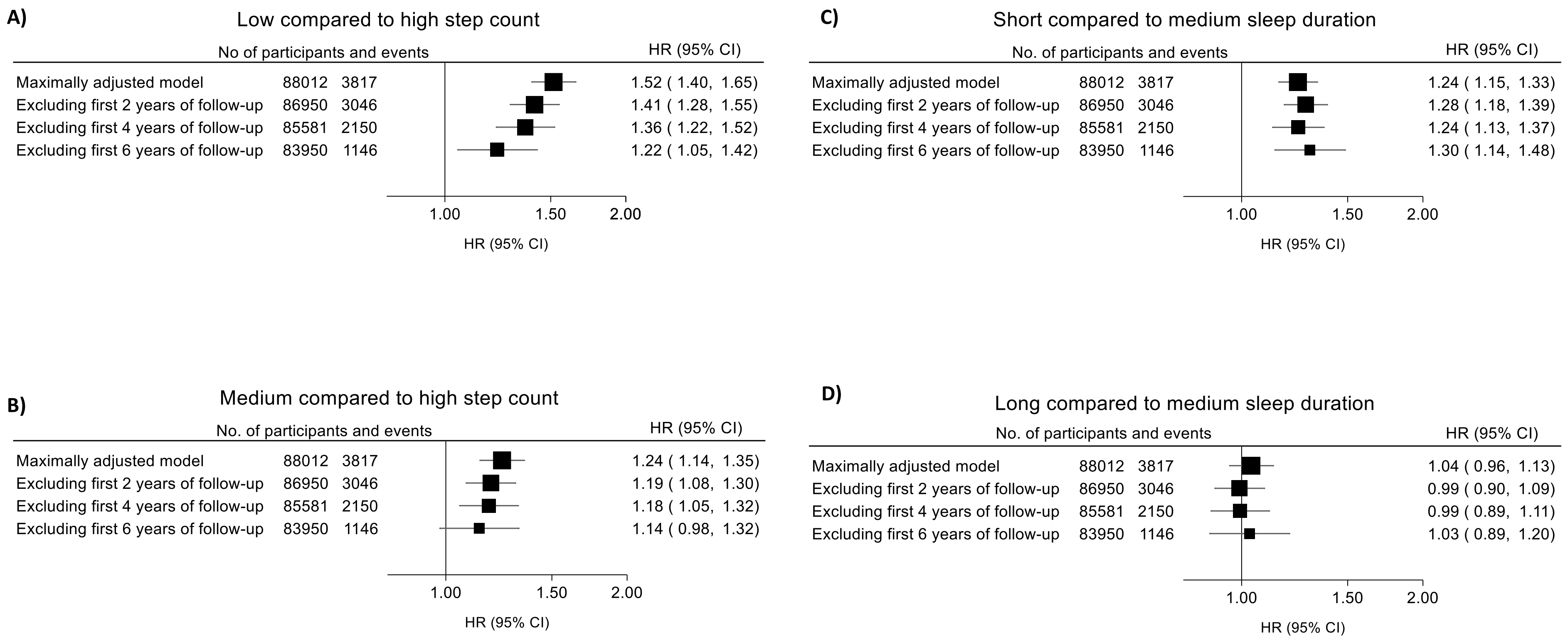


**Figure S4. Independent associations of step count (A and B) and sleep duration (C and D) with incident major adverse cardiovascular events (MACE), excluding four to six years of follow-up.**

HRs after excluding four to six years of follow-up have been depicted for each participant group according to the following classifications for median daily step count (low [<7500], intermediate [7500-11000], high [11000+; reference) and median overnight sleep duration (short [<6.5 hours], intermediate [6.5-7.5 hours; reference], long [7.5+ hours]).

The maximally adjusted model adjusted for age, sex, ethnicity, education, TDI, smoking status, alcohol intake, processed meat intake, and sleep duration or step count.

The horizontal lines represent 95% CIs based on floating absolute risks. The size of each box is relative to the amount of statistical information available. The HRs for the x-axis have been plotted on a log scale.

HR, hazard ratio; TDI, Townsend Deprivation Index; 95% CI, 95% confidence interval.

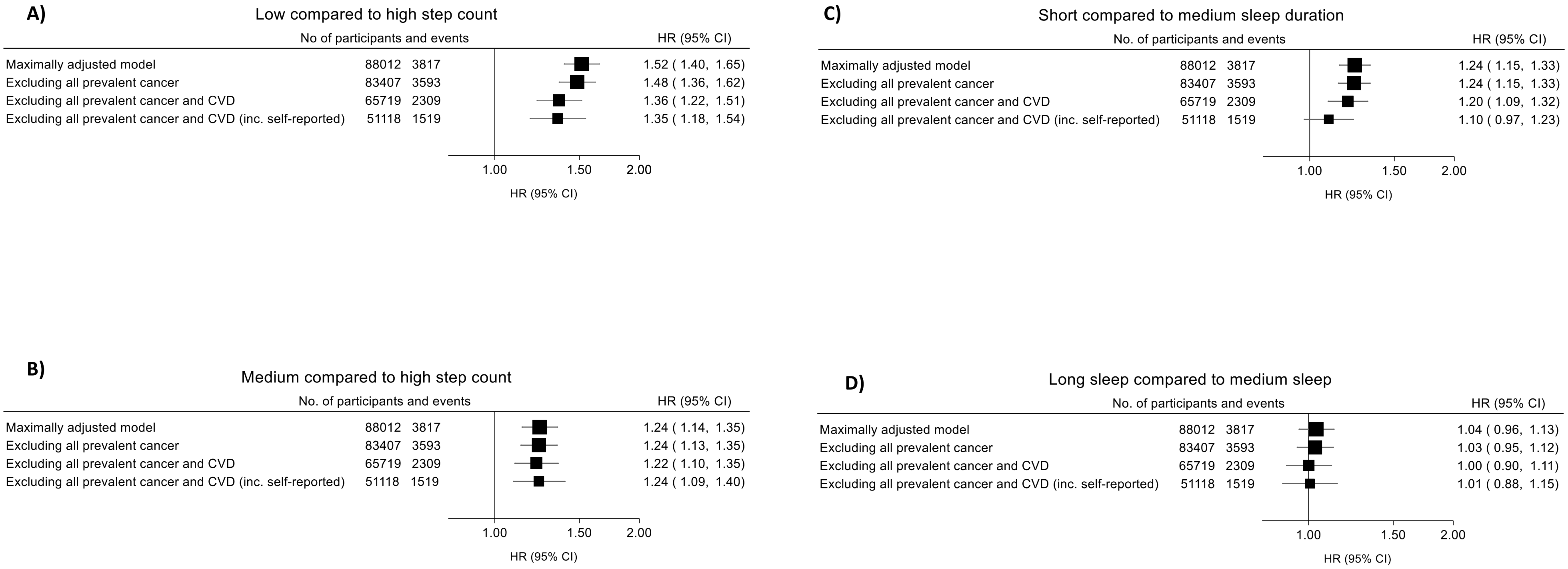


**Figure S5. Independent associations of step count (A and B) and sleep duration (C and D) with incident major adverse cardiovascular events (MACE), excluding prevalent cancer and CVD.**

HRs after sequential exclusions for prevalent cancer and CVD have been depicted for each participant group according to the following classifications for median daily step count (low [<7500], intermediate [7500-11000], high [11000+; reference) and median overnight sleep duration (short [<6.5 hours], intermediate [6.5-7.5 hours; reference], long [7.5+ hours]).

The maximally adjusted model adjusted for age, sex, ethnicity, education, TDI, smoking status, alcohol intake, processed meat intake, and sleep duration or step count.

The horizontal lines represent 95% CIs based on floating absolute risks. The size of each box is relative to the amount of statistical information available. The HRs for the x-axis have been plotted on a log scale.

CVD, cardiovascular disease; HR, hazard ratio; TDI, Townsend Deprivation Index; 95% CI, 95% confidence interval.

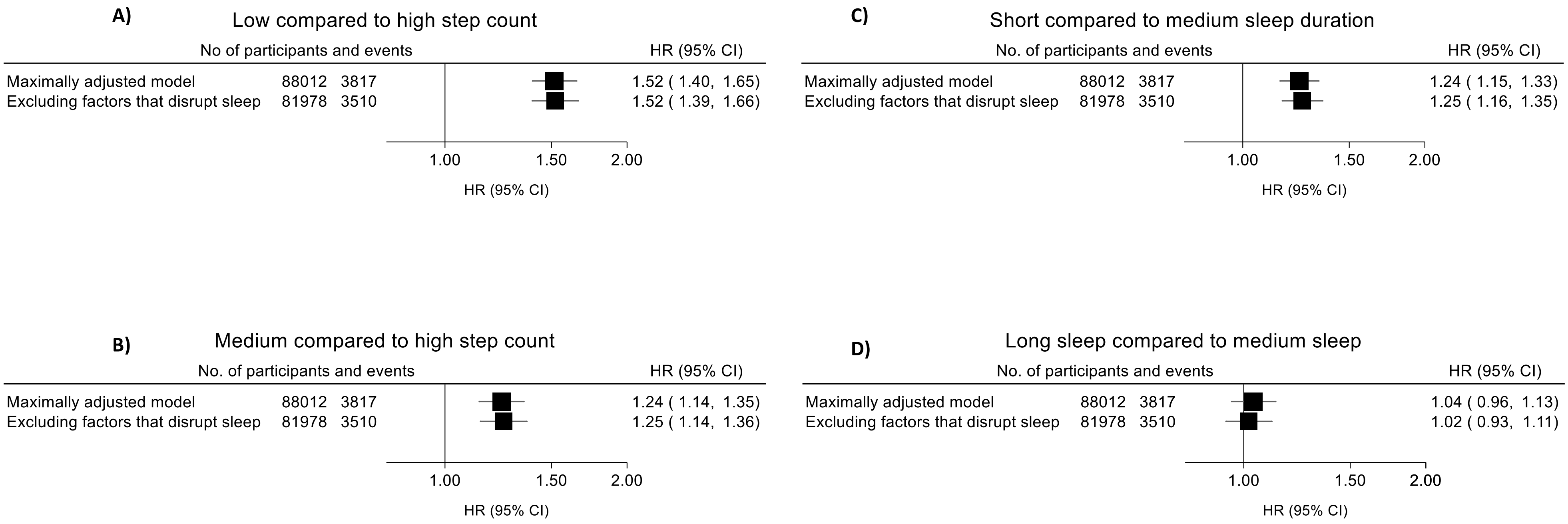


**Figure S6. Independent associations of step count (A and B) and sleep duration (C and D) with incident major adverse cardiovascular events (MACE), excluding participants with factors that disrupt sleep.**

HRs after excluding participants with factors that disrupt sleep have been depicted for each participant group according to the following classifications for median daily step count (low [<7500], intermediate [7500-11000], high [11000+; reference) and median overnight sleep duration (short [<6.5 hours], intermediate [6.5-7.5 hours; reference], long [7.5+ hours]).

Factors considered to disrupt sleep included self-reported shift work, a prior diagnosis of sleep apnoea or restless leg syndrome, and accelerometer wear that overlapped with daylight saving time changes.

The maximally adjusted model adjusted for age, sex, ethnicity, education, TDI, smoking status, alcohol intake, processed meat intake, and sleep duration or step count.

The horizontal lines represent 95% CIs based on floating absolute risks. The size of each box is relative to the amount of statistical information available. The HRs for the x-axis have been plotted on a log scale.

HR, hazard ratio; TDI, Townsend Deprivation Index; 95% CI, 95% confidence interval.

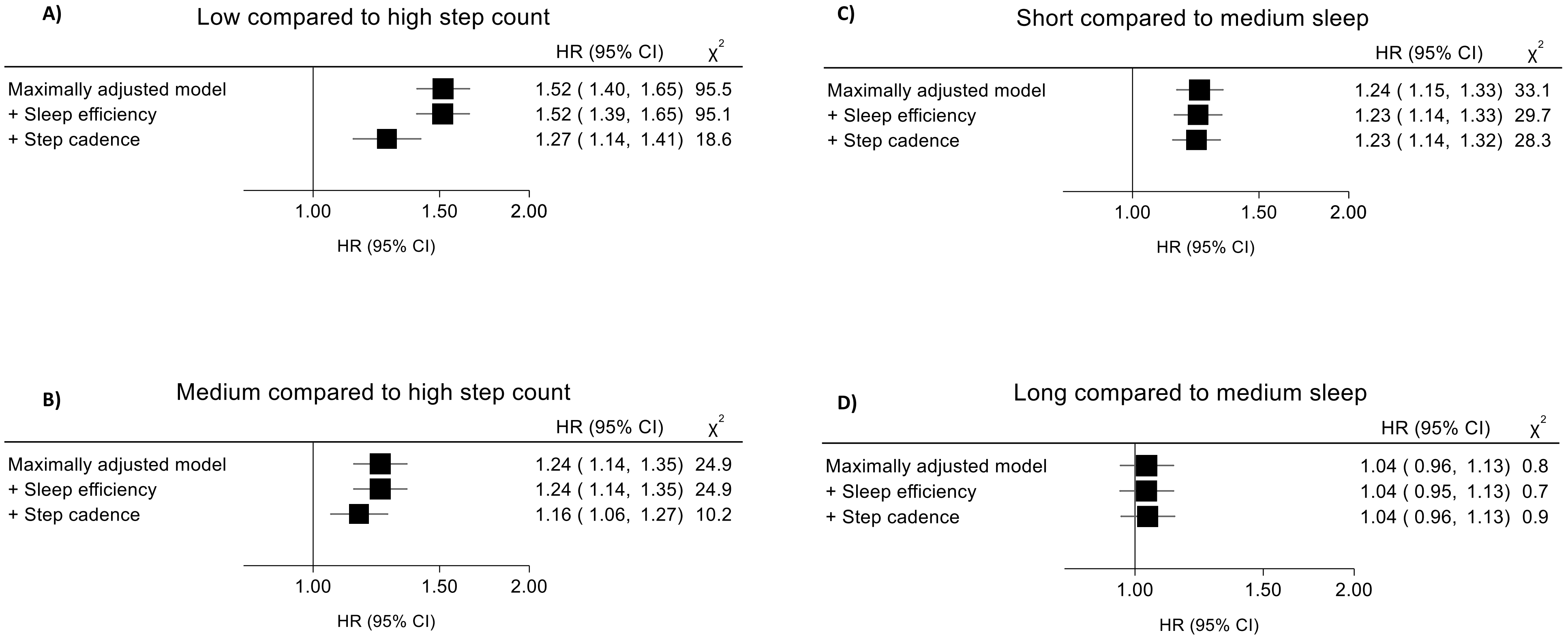


**Figure S7. Independent associations of step count (A and B) and sleep duration (C and D) with incident major adverse cardiovascular events (MACE), with sequential adjustments for sleep efficiency and step cadence.**

HRs after sequential adjustments for sleep efficiency and step cadence have been depicted for each participant group according to the following classifications for median daily step count (low [<7500], intermediate [7500-11000], high [11000+; reference) and median overnight sleep duration (short [<6.5 hours], intermediate [6.5-7.5 hours; reference], long [7.5+ hours]). $X^2$ values were derived from likelihood ratio tests comparing the maximally adjusted model with and without each covariate, and represent the potential confounding effect of that covariate. The maximally adjusted model adjusted for age, sex, ethnicity, education, TDI, smoking status, alcohol intake, processed meat intake, and sleep duration or step count.

Sleep efficiency (calculated as overnight sleep duration divided by total time in bed) and step cadence (defined as peak 30-minute cadence)[12] were both categorised into quartiles.

The horizontal lines represent 95% CIs based on floating absolute risks. The size of each box is relative to the amount of statistical information available. The HRs for the x-axis have been plotted on a log scale.

HR, hazard ratio; TDI, Townsend Deprivation Index; 95% CI, 95% confidence interval.

| | No. of participants | No. of events | HR (95% CI) |
|---|---|---|---|
| **1st Tertile** | | | |
| Short sleep duration | 7740 | 498 | 1.63 ( 1.49, 1.78) |
| Medium sleep duration | 10108 | 479 | 1.29 ( 1.18, 1.42) |
| Long sleep duration | 6738 | 377 | 1.47 ( 1.33, 1.63) |
| **2nd Tertile** | | | |
| Short sleep duration | 9104 | 447 | 1.41 ( 1.28, 1.54) |
| Medium sleep duration | 13241 | 503 | 1.16 ( 1.06, 1.27) |
| Long sleep duration | 6890 | 285 | 1.20 ( 1.07, 1.35) |
| **3rd Tertile** | | | |
| Short sleep duration | 8577 | 319 | 1.13 ( 1.01, 1.26) |
| Mediun sleep duration | 10853 | 332 | 1.00 ( 0.90, 1.11) |
| Long sleep duration | 4732 | 137 | 0.92 ( 0.78, 1.09) |

**$P_{interaction}$ = 0.54**

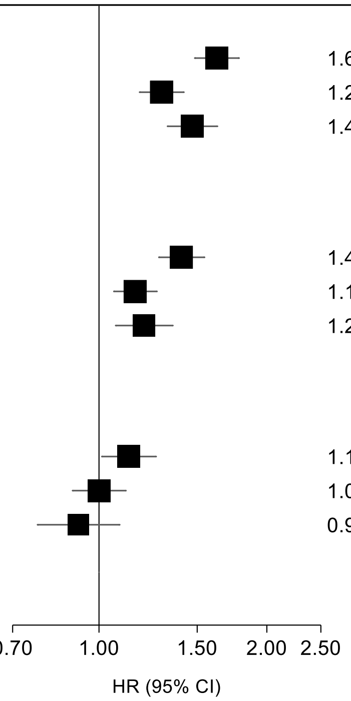


**Figure S8. Joint associations of step count and sleep duration with incident major adverse cardiovascular events (MACE), with additional adjustments for BMI, HbA1c, blood pressure, and total cholesterol.**

Additional participants were excluded due to missing covariate data, resulting in a study subpopulation of 78,866 individuals.

Participants were stratified into nine mutually exclusive groups using the following classifications for median daily step count (low [<7500], intermediate [7500-11000], high [11000+; reference) and median overnight sleep duration (short [<6.5 hours], intermediate [6.5-7.5 hours; reference], long [7.5+ hours]).

HRs after additional adjustments for potential mediators have been depicted for each participant group. The *p*-value for interaction was derived from a likelihood ratio test comparing the maximally adjusted model with and without an interaction term between step count and sleep duration. The maximally adjusted model adjusted for age, sex, ethnicity, education, TDI, smoking status, alcohol intake, and processed meat intake, then additionally BMI, HbA1c, blood pressure and total cholesterol.

For participants who self-reported taking medication for diabetes mellitus, hypertension, or hypercholesterolaemia, biomarker values were adjusted based on mean treatment effects reported in the literature.[9-11]

The horizontal lines represent 95% CIs based on floating absolute risks. The size of each box is relative to the amount of statistical information available. The HRs for the x-axis have been plotted on a log scale.

BMI, body mass index; HbA1c, glycated haemoglobin; HR, hazard ratio; TDI, Townsend Deprivation Index; 95% CI, 95% confidence interval.

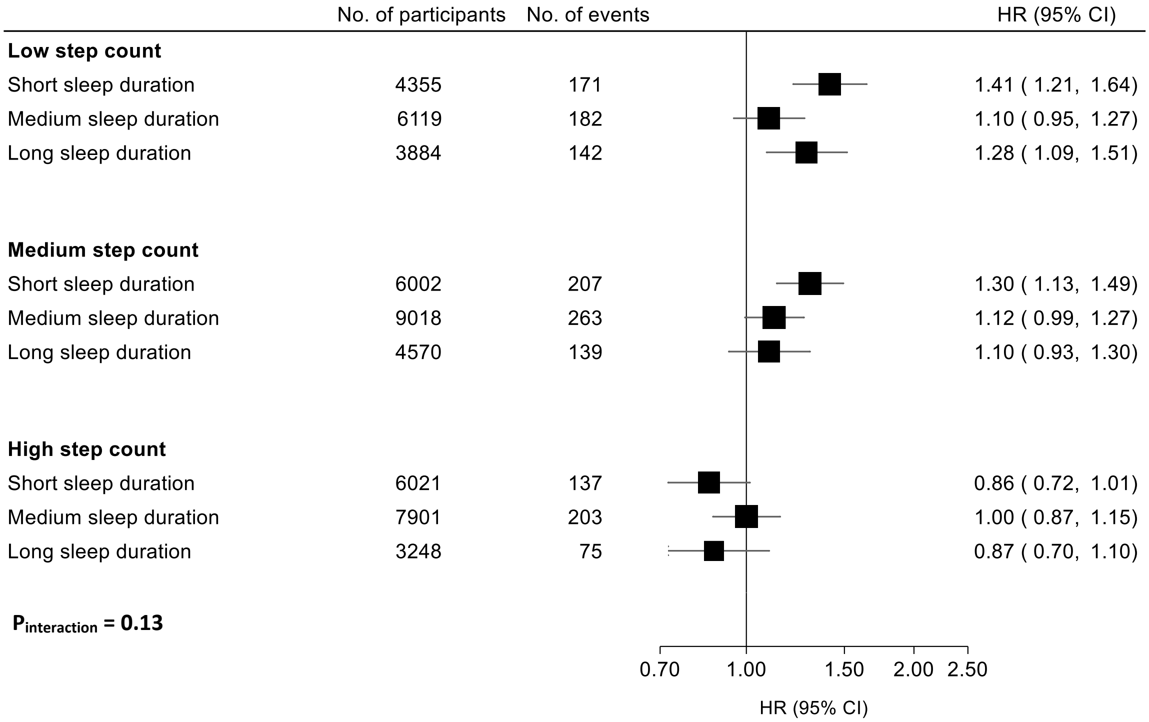


**Figure S9. Joint associations of step count and sleep duration with incident major adverse cardiovascular events (MACE), excluding prevalent cancer and CVD.**

All participants with a self-reported or hospital-recorded diagnosis of cancer or CVD were excluded, resulting in a study subpopulation of 51,118 individuals.

Participants were stratified into nine mutually exclusive groups using the following classifications for median daily step count (low [<7500], intermediate [7500-11000], high [11000+; reference) and median overnight sleep duration (short [<6.5 hours], intermediate [6.5-7.5 hours; reference], long [7.5+ hours]).

HRs after excluding prevalent cancer and CVD have been depicted for each participant group. The *p*-value for interaction was derived from a likelihood ratio test comparing the maximally adjusted model with and without an interaction term between step count and sleep duration. The maximally adjusted model adjusted for age, sex, ethnicity, education, TDI, smoking status, alcohol intake, and processed meat intake.

The horizontal lines represent 95% CIs based on floating absolute risks. The size of each box is relative to the amount of statistical information available. The HRs for the x-axis have been plotted on a log scale.

CVD, cardiovascular disease; HR, hazard ratio; TDI, Townsend Deprivation Index; 95% CI, 95% confidence interval.

| | No. of participants | No. of events | HR (95% CI) |
|---|---|---|---|
| **Low step count** | | | |
| Short sleep duration | 8456 | 325 | 1.67 ( 1.50, 1.87) |
| Medium sleep duration | 11041 | 294 | 1.21 ( 1.08, 1.36) |
| Long sleep duration | 7325 | 230 | 1.32 ( 1.16, 1.50) |
| **Medium step count** | | | |
| Short sleep duration | 9945 | 271 | 1.34 ( 1.19, 1.51) |
| Medium sleep duration | 14583 | 332 | 1.15 ( 1.03, 1.28) |
| Long sleep duration | 7540 | 182 | 1.12 ( 0.96, 1.29) |
| **High step count** | | | |
| Short sleep duration | 9453 | 207 | 1.15 ( 1.01, 1.32) |
| Medium sleep duration | 12008 | 221 | 1.00 ( 0.88, 1.14) |
| Long sleep duration | 5230 | 88 | 0.87 ( 0.71, 1.08) |

**$P_{interaction}$ = 0.44**

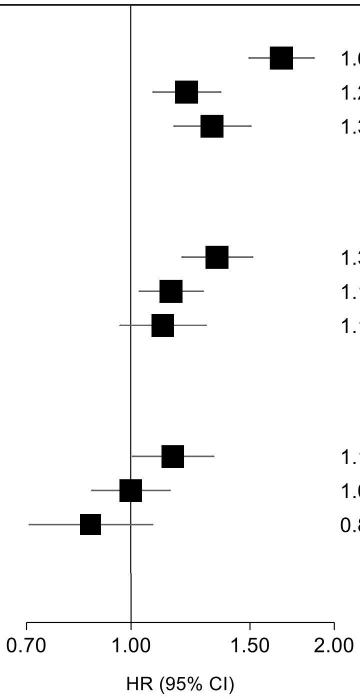


**Figure S10. Joint associations of step count and sleep duration with incident major adverse cardiovascular events (MACE), excluding four years of follow-up.**

All participants diagnosed with MACE within the first four years of follow-up were excluded, resulting in a study subpopulation of 85,581 individuals.

Participants were stratified into nine mutually exclusive groups using the following classifications for median daily step count (low [<7500], intermediate [7500-11000], high [11000+; reference) and median overnight sleep duration (short [<6.5 hours], intermediate [6.5-7.5 hours; reference], long [7.5+ hours]).

HRs after excluding four years of follow-up have been depicted for each participant group. The *p*-value for interaction was derived from a likelihood ratio test comparing the maximally adjusted model with and without an interaction term between step count and sleep duration. The maximally adjusted model adjusted for age, sex, ethnicity, education, TDI, smoking status, alcohol intake, and processed meat intake.

The horizontal lines represent 95% CIs based on floating absolute risks. The size of each box is relative to the amount of statistical information available. The HRs for the x-axis have been plotted on a log scale.

HR, hazard ratio; TDI, Townsend Deprivation Index; 95% CI, 95% confidence interval.

| | No. of participants | No. of events | HR (95% CI) |
|---|---|---|---|
| **Low step count** | | | |
| Short sleep duration | 8837 | 580 | 1.53 ( 1.40, 1.68) |
| Medium sleep duration | 11411 | 536 | 1.16 ( 1.05, 1.27) |
| Long sleep duration | 7633 | 425 | 1.31 ( 1.18, 1.45) |
| **Medium step count** | | | |
| Short sleep duration | 10261 | 496 | 1.36 ( 1.25, 1.49) |
| Medium sleep duration | 14915 | 574 | 1.13 ( 1.04, 1.23) |
| Long sleep duration | 7728 | 312 | 1.12 ( 1.00, 1.25) |
| **High step count** | | | |
| Short sleep duration | 9658 | 356 | 1.14 ( 1.02, 1.27) |
| Medium sleep duration | 12239 | 375 | 1.00 ( 0.90, 1.12) |
| Long sleep duration | 5330 | 163 | 0.97 ( 0.83, 1.14) |

**$P_{interaction}$ = 0.44**

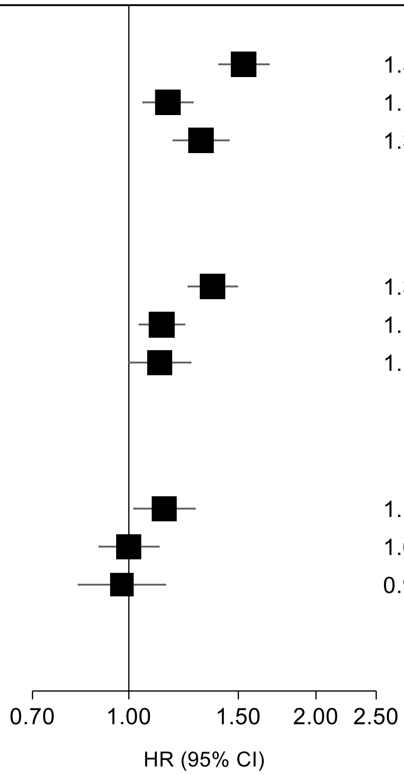


**Figure S11. Joint associations of step count and sleep duration with incident major adverse cardiovascular events (MACE), with additional adjustments for sleep efficiency and step cadence.**

Participants were stratified into nine mutually exclusive groups using the following classifications for median daily step count (low [<7500], intermediate [7500-11000], high [11000+; reference) and median overnight sleep duration (short [<6.5 hours], intermediate [6.5-7.5 hours; reference], long [7.5+ hours]).

HRs after additional adjustments for sleep efficiency and step cadence have been depicted for each participant group. The *p*-value for interaction was derived from a likelihood ratio test comparing the maximally adjusted model with and without an interaction term between step count and sleep duration. The maximally adjusted model adjusted for age, sex, ethnicity, education, TDI, smoking status, alcohol intake, and processed meat intake, then additionally sleep efficiency and step cadence.

Sleep efficiency (calculated as overnight sleep duration divided by total time in bed) and step cadence (defined as peak 30-minute cadence)[12] were both categorised into quartiles.

The horizontal lines represent 95% CIs based on floating absolute risks. The size of each box is relative to the amount of statistical information available. The HRs for the x-axis have been plotted on a log scale.

HR, hazard ratio; TDI, Townsend Deprivation Index; 95% CI, 95% confidence interval.

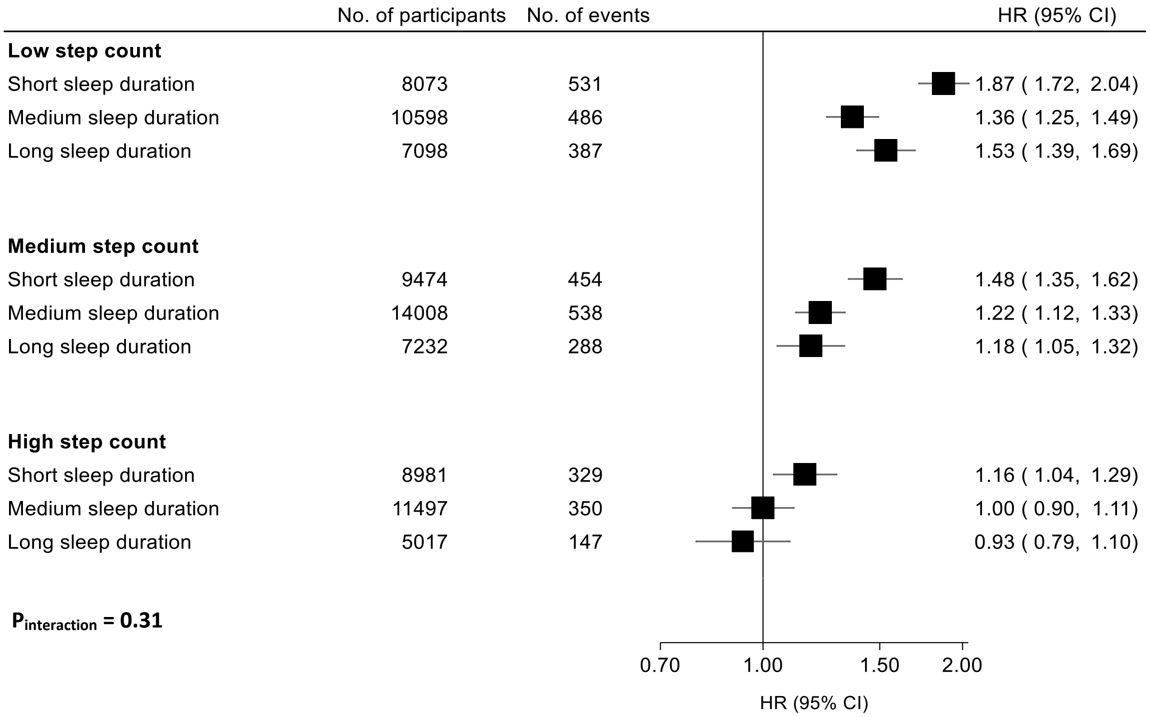


**Figure S12. Joint associations of step count and sleep duration with incident major adverse cardiovascular events (MACE), excluding participants with factors that disrupt sleep.**

Participants with self-reported shift work, a prior diagnosis of sleep apnoea or restless leg syndrome, or accelerometer wear that overlapped with daylight saving time changes were excluded, resulting in a study subpopulation of 81,987 individuals.

Participants were stratified into nine mutually exclusive groups using the following classifications for median daily step count (low [<7500], intermediate [7500-11000], high [11000+; reference) and median overnight sleep duration (short [<6.5 hours], intermediate [6.5-7.5 hours; reference], long [7.5+ hours]).

HRs after excluding participants with factors that disrupt sleep have been depicted for each participant group. The *p*-value for interaction was derived from a likelihood ratio test comparing the maximally adjusted model with and without an interaction term between step count and sleep duration. The maximally adjusted model adjusted for age, sex, ethnicity, education, TDI, smoking status, alcohol intake, and processed meat intake.

The horizontal lines represent 95% CIs based on floating absolute risks. The size of each box is relative to the amount of statistical information available. The HRs for the x-axis have been plotted on a log scale.

HR, hazard ratio; TDI, Townsend Deprivation Index; 95% CI, 95% confidence interval.

### References


1 Liang YY, Feng H, Chen Y, et al. Joint association of physical activity and sleep duration with risk of all-cause and cause-specific mortality: a population-based cohort study using accelerometry. *Eur J Prev Cardiol* 2023; **30**(9): 832-43

2 Huang BH, Duncan MJ, Cistulli PA, Nassar N, Hamer M, Stamatakis E. Sleep and physical activity in relation to all-cause, cardiovascular disease and cancer mortality risk. *Br J Sports Med* 2022; **56**(13): 718-24

3 Chen LJ, Hamer M, Lai YJ, Huang BH, Ku PW, Stamatakis E. Can physical activity eliminate the mortality risk associated with poor sleep? A 15-year follow-up of 341,248 MJ Cohort participants. *J Sport Health Sci* 2022; **11**(5): 596-604

4 Bellavia A, Åkerstedt T, Bottai M, Wolk A, Orsini N. Sleep duration and survival percentiles across categories of physical activity. *American Journal of Epidemiology* 2013; **179**(4): 484-91

5 Wang W, Yang J, Wang K, et al. Association between self-reported sleep duration, physical activity and the risk of all cause and cardiovascular diseases mortality from the NHANES database. *BMC Cardiovasc Disord* 2023; **23**(1): 467

6 Wennman H, Kronholm E, Heinonen O, et al. Leisure time physical activity and sleep predict mortality in men irrespective of background in competitive sports. *Progress in Preventive Medicine* 2017; **2**(6): e0009

7 Xiao Q, Keadle SK, Hollenbeck AR, Matthews CE. Sleep duration and total and cause-specific mortality in a large US cohort: interrelationships with physical activity, sedentary behavior, and body mass index. *Am J Epidemiol* 2014; **180**(10): 997-1006

8 Huang S, Sun H, Yu J, et al. The interaction between self-reported sleep duration and physical activity on peripheral artery disease in Chinese adults: a cross-sectional analysis in the Tianning Cohort study. *Risk Manag Healthc Policy* 2021; **14**: 4063-72

9 Canoy D, Copland E, Nazarzadeh M, et al. Antihypertensive drug effects on long-term blood pressure: an individual-level data meta-analysis of randomised clinical trials. *Heart* 2022; **108**(16): 1281-9

10 DeFronzo RA, Stonehouse AH, Han J, Wintle ME. Relationship of baseline HbA1c and efficacy of current glucose-lowering therapies: a meta-analysis of randomised clinical trials. *Diabetic Medicine* 2010; **27**(3): 309-17

11 Cholesterol Treatment Trialists' Collaboration. Efficacy and safety of LDL-lowering therapy among men and women: meta-analysis of individual data from 174,000 participants in 27 randomised trials. *The Lancet* 2015; **385**(9976): 1397-405

12 Saint-Maurice PF, Troiano RP, Bassett DR, et al. Association of Daily Step Count and Step Intensity With Mortality Among US Adults. JAMA 2020; **323**(12): 1151-60
